# Development and validation of gradient boosting decision tree models for predicting care needs using a long-term care database in Japan

**DOI:** 10.1101/2021.01.20.21250146

**Authors:** Huei-Ru Lin, Koki Fujiwara, Minoru Sasaki, Ko Ishiyama, Shino Ikeda-Sonoda, Arata Takahashi, Hiroaki Miyata

## Abstract

**Objective:** The purpose of the study was to develop machine learning models using data from long-term care (LTC) insurance claims and care needs certifications to predict the individualized future care needs of each older adult.

**Methods:** We collected LTC insurance-related data in the form of claims and care needs certification surveys from a municipality of Kanagawa Prefecture from 2009 to 2018. We used care needs certificate applications for model generation and the validation sample to build gradient boosting decision tree (GBDT) models to classify if 1) the insured’s care needs either remained stable or decreased or 2) the insured’s care needs increased after three years. The predictive model was trained and evaluated via k-fold cross-validation. The performance of the predictive model was observed in its accuracy, precision, recall, F1 score, area under the receiver-operator curve, and confusion matrix.

**Results:** Long-term care certificate applications and claim data from 2009–2018 were associated with 92,239 insureds with a mean age of 86.1 years old at the time of application, of whom 67% were female. The classifications of increase in care needs after three years were predicted with AUC of 0.80.

**Conclusions:** Machine learning is a valuable tool for predicting care needs increases in Japan’s LTC insurance system, which can be used to develop more targeted and efficient interventions to proactively reduce or prevent further functional deterioration, thereby helping older adults maintain a better quality of life.

## Introduction

The world’s population is expected to age further in the coming years. In this context, developing care systems and environments for older people is important so that they can maintain wellness in their communities as much as possible despite limited funds.^1^

Japan is among the countries with the largest aging populations as a result of its concomitant decline in birth rates and rapidly increasing proportion of population aged 65 years or older.^2^ To effectively address the challenges of an accelerating population with increased care needs, the Japan government introduced a long-term care (LTC) insurance system in 2000. The system covers all citizens aged 40 years and above.^3^ However, the LTC system eventually confronted increased financial burdens due to rapidly increasing care needs, which limited its capacity to improve quality of care in maintaining or improving the functional ability of the insured. Consequently, the Japanese government introduced an LTC prevention measurement. However, this tool has proven ineffective because of its failure to identify high-risk individuals.^4^

The administrative system in Japan consists of three levels, namely, the central government, 47 prefectures, and 1,724 municipalities.^5^ The government sets the principles and basic laws. Under the government, the Ministry of Health, Labour and Welfare is responsible for the formulation of health policies. Consequently, local government (i.e., prefectures and municipalities) embody the national health policy. Lastly, the municipalities are accountable for providing LTC services. Fifty percent of the budget for the LTC insurance is derived from general tax, whereas the other 50 percent is extracted from the premium of the insured. Thus, municipalities are insurers of the LTC insurance system.^6^

The Japanese government then asked insurers to issue analyses based on real data from LTC and medical claims to help identify insured older adults who were experiencing physical and cognitive function deterioration as well as those who were at high risk of increased care needs.^7^ The results will be an important resource that informs policies to alleviate physiological burdens of the insured and reduce financial costs for the insured and insurers.

A national standard for quantifying the care needs of the insured is called the care need level (CNL), which municipal governments use to categorize the insured on the basis of their health condition. Care needs are grouped into seven levels in ascending order. The lowest two levels are categorized as *yo-shien* (support required levels [SRLs] 1 and 2), and the higher five are denoted as *yo-kaigo* (CNLs 1 to 5).^8^ The levels define the monthly limits of the LTC insurance benefits for each insured. Individuals certified under the SRLs can only receive preventive care, such as home-based services, which is provided by municipalities. Meanwhile, the insured certified under CNLs receive home-based and institutional care services.^8^ When insured require LTC services, they submit an application to the insurers (municipalities), complete a 74-item questionnaire, and are preliminarily categorized as CNL or SRL (first certification). The applicant’s physician subsequently reviews the care level, which is finalized by an LTC insurance certification board.^9^

Previous studies have categorized the patterns and trajectories of care needs^4^ and investigated factors related to changes in care needs.^8–10^ Most of the studies have applied traditional statistical methods to develop regression models that successfully express factors related to changes in CNLs. However, they have not been able to predict the increasing care needs of each insured. Such models have been largely based on cohort datasets designed for other purposes, requiring researchers to carefully construct strategies for mitigating bias.^13^

In this context, the present study aimed to develop a model for predicting increases in the care needs of the insured. Several studies have demonstrated the usefulness of machine learning (ML) in predicting the onset of certain diseases.^14,15^ In particular, ML-related methods are suitable for predictions based on existing data,^13^ which suggests its potential applicability to the field of LTC. However, such tools have yet to be implemented. Applying ML methods to identify high-risk insured could reduce the financial burden of the insured and slow the decline in physical functioning by anticipating potential problems and implementing appropriately targeted interventions.

## Methods

The Japanese government requires insurers (municipalities) to revise health care plans every three years to ensure efficient budgeting for care benefit allocation.^16^ Therefore, the study established gradient boosting decision tree (GBDT) models to predict care needs according to the prescribed time span.

Individuals with CNL were categorized into two groups, namely, those whose CNL remained stable or decreased after three years and those whose CNL increased (Table 1). To ascertain the usefulness of the ML methods as conventional approaches, the accuracy of the model was compared to that of logistic regression under the same condition (Table A.2 in Appendix A). Additionally, we built two other ML models, one of which included four CNL categories as outcomes (i.e., declined need and loss of benefits; SRLs 1 and 2; CNL 1–3, and CNL 4–5) and another that encompassed CNL categories (i.e., the original CNL, loss, and declined need) to explore the ML models’ accuracy from different outcomes shown in Appendix B.

**Table 1.** Labels of Classification Models

| **Labels** | **Label Description** |
| --- | --- |
| 0: Stable or Improved | Care needs level either remain the same or decrease |
| 1: Care Needs Increases | Care needs level increases |

To evaluate our model, accuracies, precisions, recalls, F1 scores, area under the receiver-operator curve (AUCs), and confusion matrices of 5-fold cross validation were measured. SHapley Additive exPlanations (SHAP) was used to explain the feature importance of our model.^17^ We further explained the methods we evaluate our model refer to Appendix C.

### Dataset

The dataset was collected from the LTC database of a municipality in Kanakawa prefecture, which stores information on certificate applications and insurance claims related to care needs. In that city, the proportion of the elderly aged 65 years and above of the total population makes up about 31 percent in 2020^18^. The proportion of people certified as requiring support and care reached more than 10,000 in the end of March 2019.^19^ Structured data of 92,239 care needs certificate applications spanned from 2009 to 2018 and contained insureds’ basic information such as ID (anonymized), age, gender, application date, and certificated CNL. Each sample also came with supplemental data such as medical examination results, door-to-door surveys (74-item questionnaire of care needs certification), and LTC insurance claim data from previous years (the variables have been listed as Table A.1 in Appendix A). Because we received the dataset from the municipalities after cleaning the data to resolve privacy concerns, we could not identify the impact of missing data. Therefore, we focus on the model to establish more than the model generalization.

The predictive variable was whether an insured’s CNL increases because it represents both health conditions and expected benefits expenses of insureds. The training strategy was to use data from year *n* to predict if each insured’s care needs increased in year *n* + 3. The following two subsections described the preprocessing steps applied to the dataset.

### Data Preparation

Medical examination results and door-to-door surveys have a one-to-one relationship; thus, they are joined onto insureds’ IDs and dates of application. Insureds’ benefits have a many-to-one relationship with applications. The benefits received for a year prior to the application date were summarized as five statistical properties: sum, mean, standard deviation, and minimum and maximum values of the benefits. Each was joined with the rest of the features.

### Labeling for Target Variable

A CNL is assigned to an insured each time the insured’s application is certified. There is no record if the insured no longer applies for the certification. They may stop applying because their condition has improved, and assistance is no longer required, or they may have died or moved to another region, coming under a different insurer. For this reason, if the insured did not apply three years later, the application is removed from the dataset. Additionally, because it is impossible to predict the needs of new applicants from past application data, modeling the care needs increases of applicants that had entered the system within 1–3 years was not attempted in this study.

### Gradient Boosting Decision Tree (GBDT)

This study used GBDT models to predict the insureds’ CNL at the end of three years. GBDTs are decision-tree-based ensemble learning algorithms that are robust to multi-collinearity. More terminologies related to GBDT are provided in Appendix C.

### Training

The GBDT model was trained to predict care needs increases in three years in various granularity (Fig. 1).

**Fig 1.**
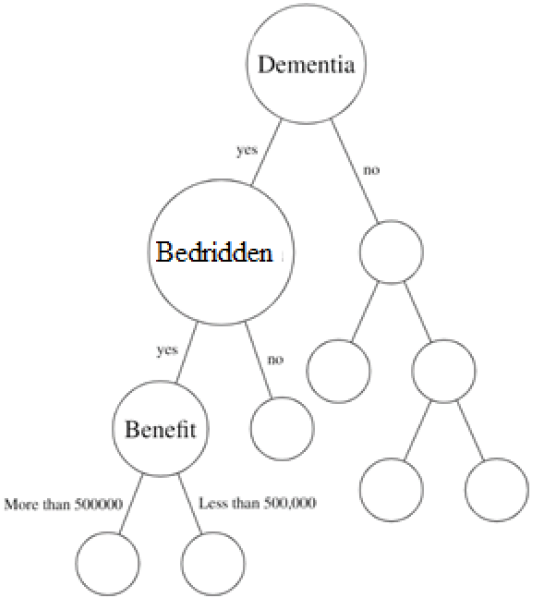
Example of a Decision Tree

Hyper-parameter optimization seeks the best hyperparameters to fit to a model. In this case, we implemented Bayesian optimization to identify the best hyperparameters to train the model.^20^ All analyses were performed using Python version 3.6.8, LightGBM version 2.2.3 for GBDT modeling and Optuna 0.10.0, for hyper-parameter optimization.

### Ethical Approval

This study was approved by the Ethics Committee of Japan Family Planning Association (ref No. 2018537).

## Results

Our results demonstrate the accuracy and overall performance of the GBDT model. A total of 92,239 care needs certificate applications in the examined 10-year period were included in our dataset. The mean age of insureds at the point of application was 86.13 years old, and 61,817 (67.0%) applicants were female. The result of statistical analysis of baseline characteristics is presented Table B.2 in Appendix B.

### Accuracy

GBDT performed better than the baseline performance. As shown in Table 2, the current model showed greater than 80% accuracy. The accuracy of the other two ML models, which included more target variable categories, increased as the number of categories decreased (see Table B.3 in Appendix B).

**Table 2.** Average Performance of Care Needs Increase Predictive Models in Three years with 5-Fold Cross-validation

| Model | Classes | Accuracy<br>(Mean ± SD), % | F1 Score<br>(Mean ± SD), % | Recall<br>(Mean ± SD), % | Precision<br>(Mean ± SD), % | AUC |
| --- | --- | --- | --- | --- | --- | --- |
| GBDT | 2 | 81 ± 1 | 80 ± 4 | 80 ± 1 | 79 ± 1 | 80 ± 1 |
| Logistic<br>Regression<sup>a</sup><br>(Base Line) | 2 | 72 ± 13 | 64 ± 11 | 63 ± 10 | 65 ± 11 | 63 ± 10 |

<sup>a</sup>The same features as the GBDT model are used for this comparison.

### AUC

The AUC is 0.80 for our model, as shown in Table 2, and the ROC curve is shown in Fig. B.3 in Appendix B. The AUC of the logistic regression was approximately 9% lower than that of the GBDT.

### Confusion Matrix

The confusion matrices shown in Fig. 2 and figs. B.1 and B.2 in Appendix B indicate that most false predictions are near the true care levels category.

**Fig 2.**
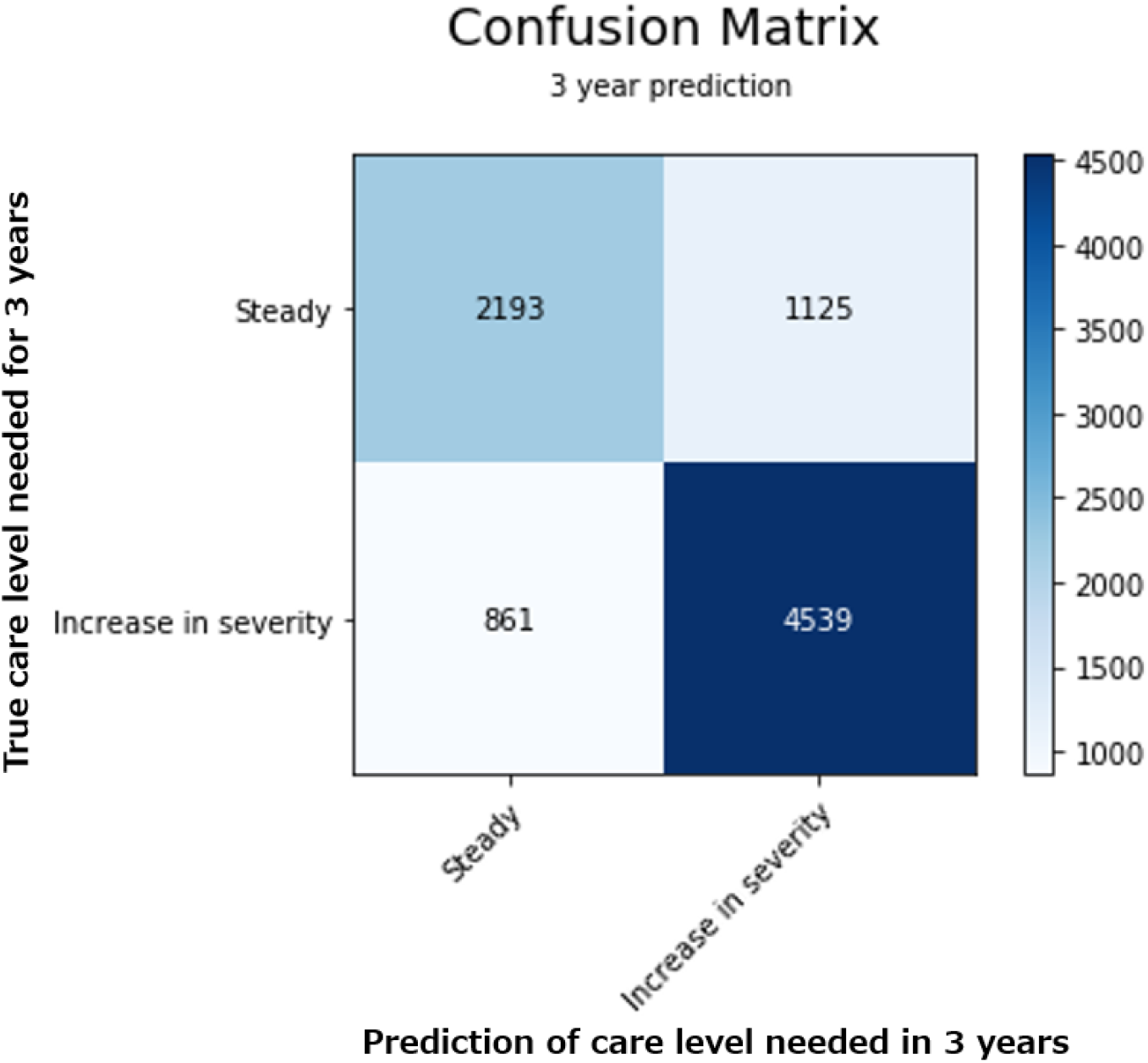
Confusion Matrix from the Two-Category Predictive Model

### Visualization

The advantage of predicting future increases in care needs at an individual level is that the model could show the probability of each individual’s increase in care needs, as well as at which CNL they will eventually be categorized. (Example shown in Fig. B.4 in Appendix B).

### Feature Importance

SHAP values were calculated to visualize the importance of explanatory variables in the binary GBDT model. The top-ranking features are provided in Fig. 3. Age exhibits the highest importance value, followed by care needs certification reference time related to dementia, intravenous therapy catheter treatment in the last two weeks, skin disease treatment in the last two weeks, insured’s CNL three years ago, and year of care needs certification. The remaining values are relatively low.

**Fig 3.**
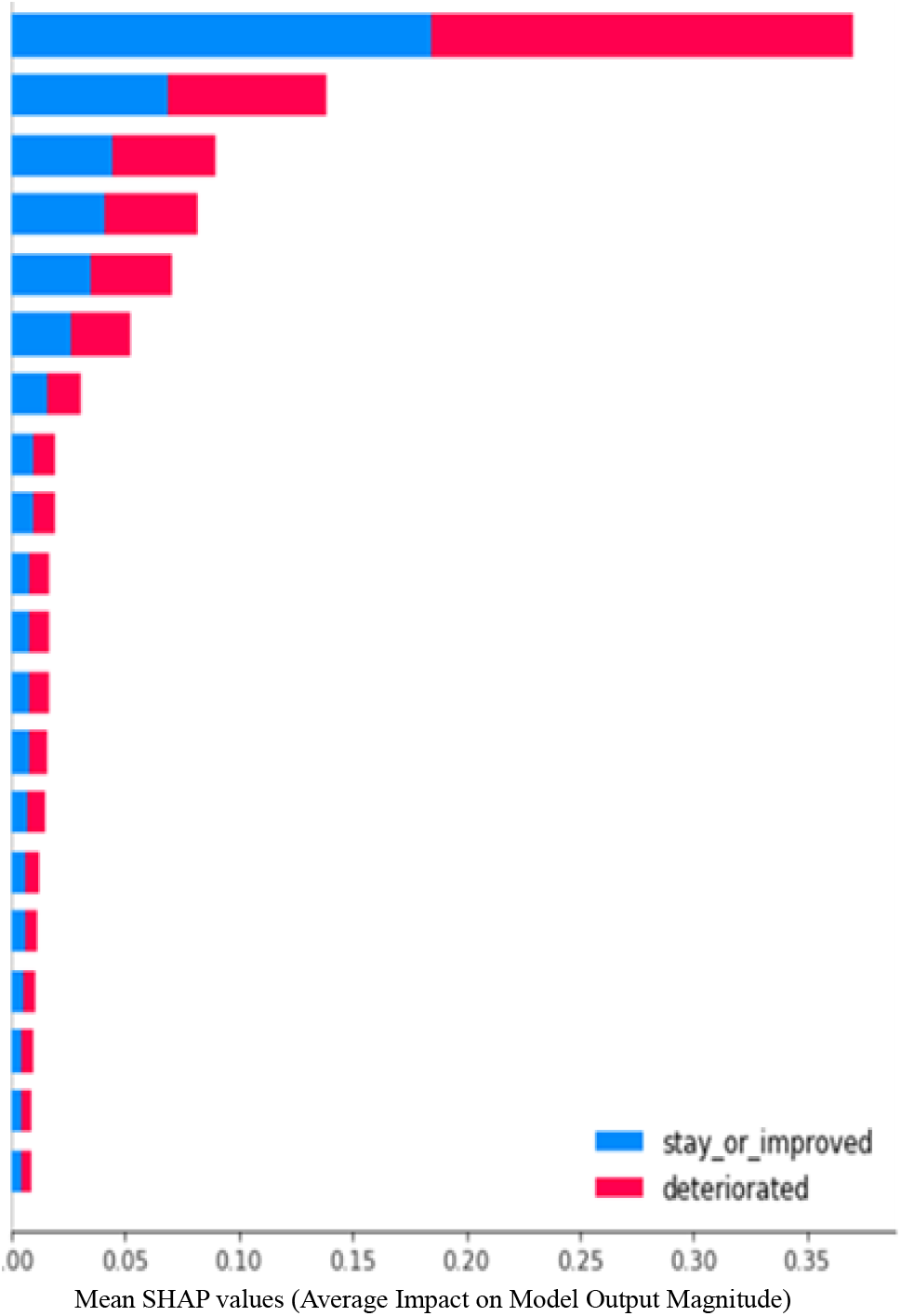
SHAP Features Importance of Binary GDBT Model

## Discussion

The purpose of the care needs increase predictive model is to help insurers foresee insureds whose health condition will deteriorate over a three-year period and thereby more effectively assess and improve the health of the insured. In comparison with previous studies, GBDT performed with better accuracy (81%, 75%, and 54% for the two-, four-, and nine-category models, respectively) than traditional regression models (72% in the two-category model) ^21,22^. The GBDT model achieved an AUC of 81%, whereas the conventional model performed at 64% (see Table B.3 of Appendix B).

The SHAP results demonstrate the logic behind the prediction. Insurers can consider and counteract the most important factors that contributed to the increases in care needs in an individual’s CNL.^23^ Consistent with previous research,^10,25–28^ this present study identified age as the most important factor related to increases in care needs. Other factors identified as important to the ML model were related to medical service use, excretion ability, daily hygiene, previous CNL, and bedridden Level 8. Among such factors, excretion ability and daily hygiene, which are activities of daily living (ADL),^28^ are the only two that may be controllable.

ADL capabilities, such as bathing, dressing, toileting, and daily hygiene, are closely related to functional and health status in older adults. Previous studies have implied that loss of independence in many ADL items is strongly associated with the use of the LTC service.^29,30^ In the current investigation, it was found that toileting and oral hygiene were the most important factors that influenced functional status in older adults. Similarly, Hirai et al.^27^ found that excretion impairment and biting ability were significantly related to an increased risk of needing LTC service. Maintaining oral health becomes difficult when individuals can no longer independently perform oral hygiene, which could result in reduced nutrient intake and increased weight loss related to oral frailty.^26,31^

The factors extracted from the LTC claims database were found to have little to no influence on increases in future care needs according to the GBDT or the logistic regression (Table A.2 in Appendix A). This result could be attributable to the fact that more than 3,000 variables were extracted from the LTC claims data. However, the majority of the variables (services) were neither used by insureds nor supplied by LTC service providers.

Fig. 2 and Appendix Figs. B.1 and B.2 show that false predictions (any off-diagonal entries) are concentrated around the diagonals. This finding indicates that although the model could not precisely predict CNL for several insured in three years, the inaccurate predictions did not deviate significantly from the categories of true needs. The study used the 74-item questionnaire for the insured to predict final CNL. However, in actuality, the finalized CNLs will be reviewed by an LTC insurance board and include data related to medical, family composition and support, and socioeconomics statuses.^32^ The lack of such additional information could explain why inaccurate predictions are concentrated around the diagonals.

CNL was not correctly predicted in approximately 23% cases within the four-category classification, among which 9% were misclassified to a higher than actual level, whereas the remaining were misclassified to some lower care needs level. However, approximately 87% of the false predictions were misclassified within a one level difference from the true class. In contrast, nearly double (45%) the proportion of cases in the nine-category classification were incorrect CNL predictions. Forty percent of the false predictions indicated a higher than actual CNL; however, 60% of the misclassifications were within a single level of the true classification.

The closer two levels are to one another, the more similar the characteristics of the insureds should be, which makes it more difficult for the models to correctly classify such their CNL. Similarly, the more resemblance between insureds at any two given levels, the greater the number of incorrect classifications between the levels. It is understandable that the boundary between care support levels is fuzzy compared to the demarcation between care support and CNL. A few insureds at higher care support levels were misclassified to CNL 1. Such individuals that have not moved to that level within three years may be more prone to do so at some later point in time. Notably, most CNL misclassifications assigned the insureds to lower rather than higher classes. A naïve explanation for this imbalance could be that some insureds might actively try to renew their care needs level for better benefits, thereby resulting in a divergence between actual and reported or perceived CNL that is reflected in higher than needed SRLs. Further analysis on those misclassified insureds may reveal some latent characteristics of insureds at each level. In addition, future work can be devoted to expanding the model’s predictive span to a five- or six-year window.

One strength of the proposed model is that it enables insurers to review the probabilities of insureds’ CNL three years in the future to glean individual trends (Appendix B). Insurers can thereby identify high-risk insureds and arrange for targeted early interventions to prevent their care needs from increasing in the near future or reduce insured’s deterioration. The SHAP analysis identifies most important factors impacting each insured’s CNL, which is highly informative for determining the most effective interventions. Overall, the model could assist insurers to improve targeted decision-making to implement services more efficiently and reduce wasteful costs.^7^

CNL 5 was the highest predicted level in this study, which is likely because our model is restricted to a three-year window. Moreover, some applicants are likely to become deceased and be removed from the records. The records do not differentiate such cases from other situations of data loss such as losing track of applicants due to moving events.

To the best of our knowledge, this is the first model that predicts the CNL increases of insured individuals using the LTC database (including claims and CNL certification data). The ability to predict and clarify the most important factors related to the future care needs increases of each insured can help insurers determine the appropriate interventions to limit or prevent increasing care needs, thereby reducing the financial burdens of both the LTC system and the patients. Most importantly, keeping patients’ care needs at a stable level corresponds to slower declines in physical or cognitive function and thus promotes better quality of life for the elderly. These results are useful for helping policymakers and providers to narrow their focus from the currently broad classifications and devise more targeted and efficient measures informed by the specific factors that impact different insured individuals.

### Limitations and Future Research

In this research, we established three models utilizing a large-scale sample of older adults living in one Japanese city over a period 10 years. A large sample is beneficial because it leads to more stable results and a higher possibility of generalizability. However, the situation of insureds may change with time, and this effect needs to be considered in future research. In addition, the LTC database could express only part of the situations of insureds. To increase the model’s predictability, additional data such as health check-up results and other medical records, social capital, living arrangements, socioeconomic status, and family support (informal care) are needed in order to comprehensively clarify the conditions and needs of older adults.

Regarding the privacy concerns, we received the dataset after the municipality conducted data cleaning; therefore, we were unable to identify the impact that resulted from data that were excluded or censored for multiple reasons. Furthermore, the insureds who applied the care needs certification for the first time and those who had no data in the previous year were excluded from our analysis. These may therefore affect the generalizability of our findings.

Future research should not only explore rich data but also explain the impact of the data that is missing to better extend the model’s generalization.

## Conclusion

This study built a ML model that was able to predict the CNL of insured individuals with greater accuracy than other methods by using LTC database of records compiled over periods spanning three years. The results indicate that ML models can be used to 1) identify individuals at high risk of increased care needs and 2) represent targeted risk factors related to care needs changes. These results could prevent care needs increases by providing information relevant to designing and implementing early interventions, thereby reducing care financial burdens and maintaining the insured’s functional ability and promoting better quality of life.

## Supporting information

Appendix A Results of the logistic regression model

Appendix B Outcomes of the machine learning models

Appendix C Evaluated methods

## Data Availability

The datasets generated and analyzed during the current study are available the corresponding author on resonable request. Please contact the Ethics Committee, Japan　Family Planning Association，Inc. and data owner Kanakawa prefecture MeByo labo (http://www.pref.kanagawa.jp/docs/bs5/cnt/f536534/index.html) for further information regarding the data availability of this study.

## Acknowledgments

This study used data from the Kanagawa Prefecture ME-BYO Living Lab. The authors thank Prof. Miya Kobayashi, all staff members of the ME-BYO Living Lab in Kanagawa Prefecture, and Masayuki Nakane for providing valuable suggestions.

