## Appendix A Results of the logistic regression model for "Development and validation of gradient boosting decision tree models for predicting care needs using a long-term care database in Japan"

Results of the logistic regression model to clarify the factors related to increase in care needs of long-term care insureds in Japan in 3 years.

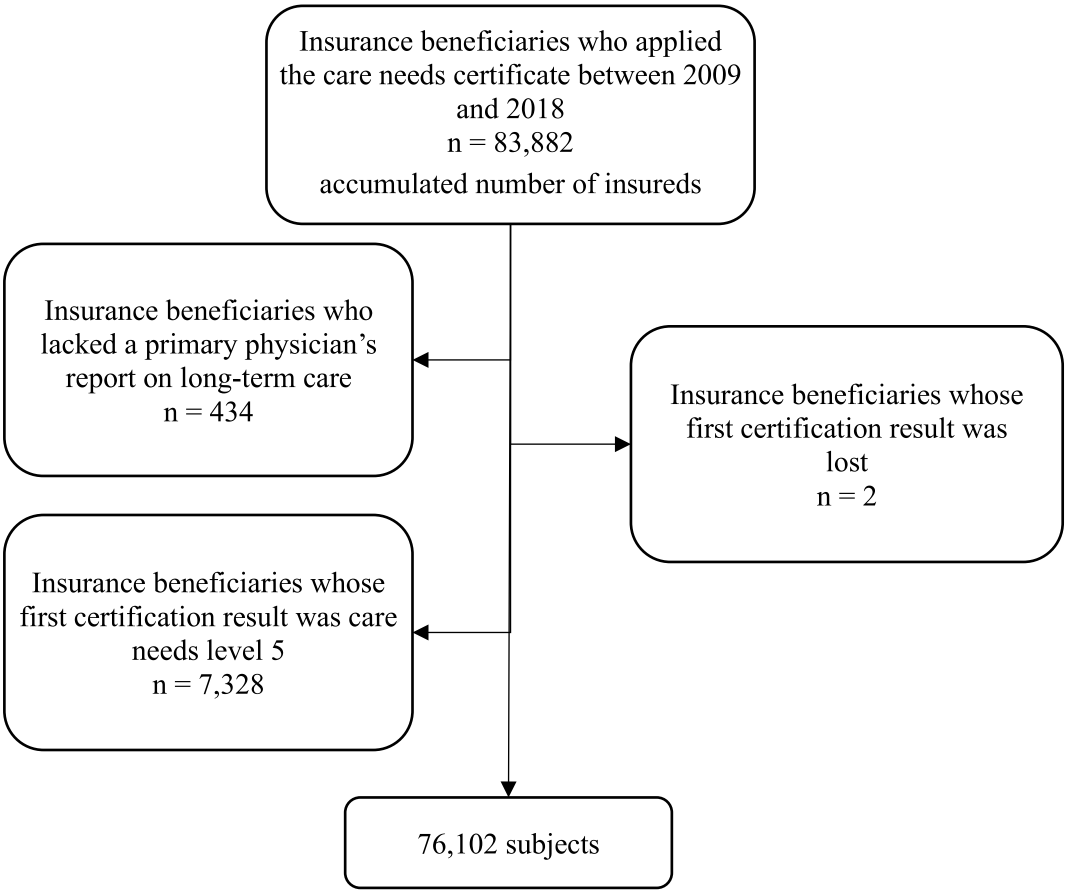


Fig. A.1. Flow chart of data selection; the study sample for analysis comprised 76,102 subjects.

Table A.1.

Demographic Characteristics and Chi-Square Test Result of Numbers of Stable or Improved and Deteriorated Groups.

| Variables | Stable or Improved (care needs remained the same or decreased) n = 35,289 | Deteriorated (care needs increased) n = 40,813 | Total n = 76,102 | Deterioration Proportion (%) | % in each category | P value* |
|---|---|---|---|---|---|---|
| Sex | | | | | | <0.0001 |
| Male | 10,879 | 13,835 | 24,714 | 56.0% | 32.5% | |
| Female | 24,410 | 26,978 | 51,388 | 52.5% | 67.5% | |
| Former care needs certification result_1st judgment | | | | | | <0.0001 |
| Unknown | 6,100 | 8,298 | 14,398 | 57.6% | 18.9% | |
| Unqualified | 718 | 1,095 | 1,813 | 60.4% | 2.4% | |
| Temporary long-term care level | 303 | 512 | 815 | 62.8% | 1.1% | |
| Support required level 1 | 5,799 | 5,974 | 11,773 | 50.7% | 15.5% | |
| Support required level 2 | 5,606 | 6,332 | 11,938 | 53.0% | 15.7% | |
| Care needs level 1 | 6,660 | 8,497 | 15,157 | 56.1% | 19.9% | |
| Care needs level 2 | 4,788 | 5,129 | 9,917 | 51.7% | 13.0% | |
| Care needs level 3 | 2,759 | 2,249 | 5,008 | 44.9% | 6.6% | |
| Care needs level 4 | 2,086 | 1,866 | 3,952 | 47.2% | 5.2% | |
| Care needs level 5 | 470 | 861 | 1,331 | 64.7% | 1.7% | |
| Former care needs certification result_2nds judgment | | | | | | <0.0001 |
| Unknown | 5,781 | 7,926 | 13,707 | 57.8% | 18.0% | |
| Unqualified | 224 | 343 | 567 | 60.5% | 0.7% | |

| Variables | Stable or Improved (care needs remained the same or decreased) | Deteriorated (care needs increased) | Total | Deterioration Proportion (%) | % in each category | P value* |
|---|---|---|---|---|---|---|
| | n = 35,289 | n = 40,813 | n = 76,102 | | | |
| Temporary long-term care level | 55 | 80 | 135 | 59.3% | 0.2% | |
| Support required level 1 | 5,896 | 5,918 | 11,814 | 50.1% | 15.5% | |
| Support required level 2 | 5,385 | 5,753 | 11,138 | 51.7% | 14.6% | |
| Care needs level 1 | 6,741 | 8,257 | 14,998 | 55.1% | 19.7% | |
| Care needs level 2 | 4,997 | 5,803 | 10,800 | 53.7% | 14.2% | |
| Care needs level 3 | 3,379 | 3,570 | 6,949 | 51.4% | 9.1% | |
| care needs level 4 | 2,389 | 2,043 | 4,432 | 46.1% | 5.8% | |
| Care needs level 5 | 442 | 1,120 | 1,562 | 71.7% | 2.1% | |
| Living arrangement | | | | | | 0.076 |
| Unknown | 2 | 0 | 2 | 0.0% | 0.0% | |
| Home | 34,332 | 39,788 | 74,120 | 53.7% | 97.4% | |
| Facility | 955 | 1,025 | 1,980 | 51.8% | 2.6% | |
| Care Needs Level_1st judgment | | | | | | <0.0001 |
| Unqualified | 16 | 1,799 | 1,815 | 99.1% | 2.4% | |
| Support required level 1 | 5,572 | 8,290 | 13,862 | 59.8% | 18.2% | |
| Support required level 2 | 6,518 | 7,467 | 13,985 | 53.4% | 18.4% | |
| Care needs level 1 | 6,827 | 9,914 | 16,741 | 59.2% | 22.0% | |
| Care needs level 2 | 6,593 | 6,638 | 13,231 | 50.2% | 17.4% | |
| Care needs level 3 | 4,210 | 3,532 | 7,742 | 45.6% | 10.2% | |

| Variables | Stable or Improved (care needs remained the same or decreased) | Deteriorated (care needs increased) | Total | Deterioration Proportion (%) | % in each category | P value* |
|---|---|---|---|---|---|---|
| | n = 35,289 | n = 40,813 | n = 76,102 | | | |
| Care needs level 4 | 5,553 | 3,173 | 8,726 | 36.4% | 11.5% | |
| Care Needs Level_2nd judgment | | | | | | <0.0001 |
| Unqualified | 41 | 366 | 407 | 89.9% | 0.5% | |
| Support required level 1 | 5,596 | 7,595 | 13,191 | 57.6% | 17.3% | |
| Support required level 2 | 6,199 | 5,876 | 12,075 | 48.7% | 15.9% | |
| Care needs level 1 | 7,035 | 10,049 | 17,084 | 58.8% | 22.4% | |
| Care needs level 2 | 6,531 | 7,079 | 13,610 | 52.0% | 17.9% | |
| Care needs level 3 | 4,389 | 5,433 | 9,822 | 55.3% | 12.9% | |
| Care needs level 4 | 5,439 | 3,287 | 8,726 | 37.7% | 11.5% | |
| Care needs level 5 | 59 | 1,128 | 1,187 | 95.0% | 1.6% | |
| Degree of independent living for elderly with disability | | | | | | <0.0001 |
| Dependent | 164 | 274 | 438 | 62.6% | 0.6% | |
| Ｊ１（able to go out using public transportation） | 3,514 | 3,344 | 6,858 | 48.8% | 9.0% | |
| Ｊ２（able to go out but limited to the neighborhood） | 6,999 | 8,186 | 15,185 | 53.9% | 20.0% | |
| Ａ１（able to go out with assistance and usually not bedridden） | 7,195 | 9,927 | 17,122 | 58.0% | 22.5% | |
| Ａ２（rarely able to go out and naps several times during daytime） | 8,363 | 11,249 | 19,612 | 57.4% | 25.8% | |

| Variables | Stable or Improved (care needs remained the same or decreased) n = 35,289 | Deteriorated (care needs increased) n = 40,813 | Total n = 76,102 | Deterioration Proportion (%) | % in each category | P value* |
|---|---|---|---|---|---|---|
| Ｂ１（able to eat and use a wheelchair to access the toilet away from bed） | 3,491 | 3,442 | 6,933 | 49.6% | 9.1% | |
| Ｂ２（requires care to get on a wheelchair） | 4,222 | 2,729 | 6,951 | 39.3% | 9.1% | |
| Ｃ１（able to roll over by him/herself） | 794 | 612 | 1,406 | 43.5% | 1.8% | |
| Ｃ２（fully assisted in rolling over by him/herself） | 547 | 1,050 | 1,597 | 65.7% | 2.1% | |
| Degree of independent living for elderly people with dementia | | | | | | <0.0001 |
| Dependent | 9,478 | 9,365 | 18,843 | 49.7% | 24.8% | |
| I | 9,917 | 11,536 | 21,453 | 53.8% | 28.2% | |
| II a | 3,902 | 4,279 | 8,181 | 52.3% | 10.8% | |
| II b | 6,495 | 7,221 | 13,716 | 52.6% | 18.0% | |
| III a | 3,775 | 5,450 | 9,225 | 59.1% | 12.1% | |
| III b | 869 | 1,251 | 2,120 | 59.0% | 2.8% | |
| IV | 759 | 1,442 | 2,201 | 65.5% | 2.9% | |
| Ｍ | 94 | 269 | 363 | 74.1% | 0.5% | |
| Short-term memory | | | | | | <0.0001 |
| Loss | 12,432 | 12,864 | 25,296 | 50.9% | 33.2% | |
| Unknown | 11 | 3 | 14 | 21.4% | 0.0% | |
| Independent | 11,066 | 11,962 | 23,028 | 51.9% | 30.3% | |
| With difficulty | 11,780 | 15,984 | 27,764 | 57.6% | 36.5% | |

| Variables | Stable or Improved (care needs remained the same or decreased) | Deteriorated (care needs increased) | Total | Deterioration Proportion (%) | % in each category | P value* |
|---|---|---|---|---|---|---|
| | n = 35,289 | n = 40,813 | n = 76,102 | | | |
| Cognitive ability (daily decision-making) | | | | | | <0.0001 |
| Loss | 12,432 | 12,864 | 25,296 | 50.9% | 33.2% | |
| Unknown | 13 | 5 | 18 | 27.8% | 0.0% | |
| Independent | 11,208 | 12,289 | 23,497 | 52.3% | 30.9% | |
| Some difficulty | 6,482 | 8,183 | 14,665 | 55.8% | 19.3% | |
| Partially assisted | 3,903 | 5,374 | 9,277 | 57.9% | 12.2% | |
| Fully assisted | 1,251 | 2,098 | 3,349 | 62.6% | 4.4% | |
| Communication ability | | | | | | <0.0001 |
| Unknown | 14 | 7 | 21 | 33.3% | 0.0% | |
| Capable | 13,528 | 15,312 | 28,840 | 53.1% | 37.9% | |
| Somewhat challenging | 5,876 | 7,572 | 13,448 | 56.3% | 17.7% | |
| Limited to specific requirements | 2,953 | 4,050 | 7,003 | 57.8% | 9.2% | |
| Incapable | 486 | 1,008 | 1,494 | 67.5% | 2.0% | |
| Eating | | | | | | <0.0001 |
| Unknown | 17 | 25 | 42 | 59.5% | 0.1% | |
| Independent | 22,326 | 26,937 | 49,263 | 54.7% | 64.7% | |
| Needs to be fed | 514 | 987 | 1,501 | 65.8% | 2.0% | |
| BPSD | | | | | | <0.0001 |
| None | 5,225 | 5,686 | 10,911 | 52.1% | 14.3% | |

| Variables | Stable or Improved (care needs remained the same or decreased) | Deteriorated (care needs increased) | Total | Deterioration Proportion (%) | % in each category | P value* |
|---|---|---|---|---|---|---|
| | n = 35,289 | n = 40,813 | n = 76,102 | | | |
| Yes | 3,272 | 5,204 | 8,476 | 61.4% | 11.1% | |
| No | 14,360 | 17,059 | 31,419 | 54.3% | 41.3% | |
| Turn in bed | | | | | | <0.0001 |
| Independent | 11,742 | 17,824 | 29,566 | 60.3% | 38.9% | |
| Can do holding railings | 20,575 | 20,069 | 40,644 | 49.4% | 53.4% | |
| Fully assisted | 2,972 | 2,920 | 5,892 | 49.6% | 7.7% | |
| Get up | | | | | | <0.0001 |
| Independent | 2,738 | 4,963 | 7,701 | 64.4% | 10.1% | |
| Can do holding railings | 28,633 | 32,345 | 60,978 | 53.0% | 80.1% | |
| Fully assisted | 3,918 | 3,505 | 7,423 | 47.2% | 9.8% | |
| Maintain sitting position | | | | | | <0.0001 |
| Independent | 12,519 | 17,827 | 30,346 | 58.7% | 39.9% | |
| Can do holding railings | 13,864 | 15,032 | 28,896 | 52.0% | 38.0% | |
| Partially assisted | 8,626 | 7,312 | 15,938 | 45.9% | 20.9% | |
| Fully assisted | 280 | 642 | 922 | 69.6% | 1.2% | |
| Stand on both feet | | | | | | <0.0001 |
| Independent | 15,376 | 21,239 | 36,615 | 58.0% | 48.1% | |
| Partially assisted | 16,690 | 16,636 | 33,326 | 49.9% | 43.8% | |
| Fully assisted | 3,223 | 2,938 | 6,161 | 47.7% | 8.1% | |

| Variables | Stable or Improved (care needs remained the same or decreased) | Deteriorated (care needs increased) | Total | Deterioration Proportion (%) | % in each category | P value* |
|---|---|---|---|---|---|---|
| | n = 35,289 | n = 40,813 | n = 76,102 | | | |
| Walk | | | | | | <0.0001 |
| Independent | 8,718 | 12,402 | 21,120 | 58.7% | 27.8% | |
| Can do with assistive technology | 19,301 | 22,299 | 41,600 | 53.6% | 54.7% | |
| Fully assisted | 7,270 | 6,112 | 13,382 | 45.7% | 17.6% | |
| Transfer | | | | | | <0.0001 |
| Independent | 20,631 | 27,936 | 48,567 | 57.5% | 63.8% | |
| Needs supervision | 6,700 | 7,025 | 13,725 | 51.2% | 18.0% | |
| Partially assisted | 5,514 | 3,523 | 9,037 | 39.0% | 11.9% | |
| Fully assisted | 2,444 | 2,329 | 4,773 | 48.8% | 6.3% | |
| Stand up | | | | | | <0.0001 |
| Independent | 1,027 | 2,955 | 3,982 | 74.2% | 5.2% | |
| Can do holding railings | 30,411 | 34,545 | 64,956 | 53.2% | 85.4% | |
| Fully assisted | 3,851 | 3,313 | 7,164 | 46.2% | 9.4% | |
| Stand on one foot | | | | | | <0.0001 |
| Independent | 1,909 | 3,369 | 5,278 | 63.8% | 6.9% | |
| Partially assisted | 21,312 | 26,671 | 47,983 | 55.6% | 63.1% | |
| Fully assisted | 12,068 | 10,773 | 22,841 | 47.2% | 30.0% | |
| Bathing | | | | | | <0.0001 |
| Independent | 13,659 | 16,599 | 30,258 | 54.9% | 39.8% | |

| Variables | Stable or Improved (care needs remained the same or decreased) | Deteriorated (care needs increased) | Total | Deterioration Proportion (%) | % in each category | P value* |
|---|---|---|---|---|---|---|
| | n = 35,289 | n = 40,813 | n = 76,102 | | | |
| Partially assisted | 11,511 | 13,982 | 25,493 | 54.8% | 33.5% | |
| Fully assisted | 7,984 | 7,793 | 15,777 | 49.4% | 20.7% | |
| Does not take bath | 2,135 | 2,439 | 4,574 | 53.3% | 6.0% | |
| Swallowing | | | | | | <0.0001 |
| Independent | 26,431 | 32,093 | 58,524 | 54.8% | 76.9% | |
| Needs supervision | 8,635 | 7,888 | 16,523 | 47.7% | 21.7% | |
| Fully assisted | 223 | 832 | 1,055 | 78.9% | 1.4% | |
| Eating | | | | | | <0.0001 |
| Independent | 28,818 | 33,982 | 62,800 | 54.1% | 82.5% | |
| Needs supervision | 4,146 | 4,439 | 8,585 | 51.7% | 11.3% | |
| Partially assisted | 1,969 | 1,394 | 3,363 | 41.5% | 4.4% | |
| Fully assisted | 356 | 998 | 1,354 | 73.7% | 1.8% | |
| Oral hygiene | | | | | | <0.0001 |
| Independent | 23,642 | 28,750 | 52,392 | 54.9% | 68.8% | |
| Partially assisted | 9,545 | 9,580 | 19,125 | 50.1% | 25.1% | |
| Fully assisted | 2,102 | 2,483 | 4,585 | 54.2% | 6.0% | |
| Washing face | | | | | | <0.0001 |
| Independent | 23,568 | 29,054 | 52,622 | 55.2% | 69.1% | |
| Partially assisted | 9,623 | 9,292 | 18,915 | 49.1% | 24.9% | |

| Variables | Stable or Improved (care needs remained the same or decreased) | Deteriorated (care needs increased) | Total | Deterioration Proportion (%) | % in each category | P value* |
|---|---|---|---|---|---|---|
| | n = 35,289 | n = 40,813 | n = 76,102 | | | |
| Fully assisted | 2,098 | 2,467 | 4,565 | 54.0% | 6.0% | |
| Hair care | | | | | | <0.0001 |
| Independent | 25,800 | 31,066 | 56,866 | 54.6% | 74.7% | |
| Partially assisted | 6,004 | 6,057 | 12,061 | 50.2% | 15.8% | |
| Fully assisted | 3,485 | 3,690 | 7,175 | 51.4% | 9.4% | |
| Nail-cutting | | | | | | <0.0001 |
| Independent | 13,044 | 16,303 | 29,347 | 55.6% | 38.6% | |
| Partially assisted | 8,910 | 10,085 | 18,995 | 53.1% | 25.0% | |
| Fully assisted | 13,335 | 14,425 | 27,760 | 52.0% | 36.5% | |
| Putting on/removing top | | | | | | <0.0001 |
| Independent | 19,826 | 24,869 | 44,695 | 55.6% | 58.7% | |
| Needs supervision | 2,795 | 4,673 | 7,468 | 62.6% | 9.8% | |
| Partially assisted | 9,783 | 8,444 | 18,227 | 46.3% | 24.0% | |
| Fully assisted | 2,885 | 2,827 | 5,712 | 49.5% | 7.5% | |
| Putting on/removing pants | | | | | | <0.0001 |
| Independent | 18,836 | 24,133 | 42,969 | 56.2% | 56.5% | |
| Needs supervision | 2,739 | 4,487 | 7,226 | 62.1% | 9.5% | |
| Partially assisted | 8,805 | 8,109 | 16,914 | 47.9% | 22.2% | |
| Fully assisted | 4,909 | 4,084 | 8,993 | 45.4% | 11.8% | |

| Variables | Stable or Improved (care needs remained the same or decreased) | Deteriorated (care needs increased) | Total | Deterioration Proportion (%) | % in each category | P value* |
|---|---|---|---|---|---|---|
| | n = 35,289 | n = 40,813 | n = 76,102 | | | |
| Taking medication | | | | | | 0.048 |
| Independent | 14,734 | 16,962 | 31,696 | 53.5% | 41.6% | |
| Partially assisted | 15,489 | 18,206 | 33,695 | 54.0% | 44.3% | |
| Fully assisted | 5,066 | 5,645 | 10,711 | 52.7% | 14.1% | |
| Managing money | | | | | | <0.0001 |
| Independent | 12,791 | 14,261 | 27,052 | 52.7% | 35.5% | |
| Partially assisted | 7,320 | 8,957 | 16,277 | 55.0% | 21.4% | |
| Fully assisted | 15,178 | 17,595 | 32,773 | 53.7% | 43.1% | |
| Vision | | | | | | <0.0001 |
| Normal | 23,788 | 28,364 | 52,152 | 54.4% | 68.5% | |
| Able to see over 1m away | 9,180 | 9,669 | 18,849 | 51.3% | 24.8% | |
| Nearsightedness | 1,726 | 1,794 | 3,520 | 51.0% | 4.6% | |
| Poor sight | 434 | 472 | 906 | 52.1% | 1.2% | |
| Cannot be judged | 161 | 514 | 675 | 76.1% | 0.9% | |
| Hearing ability | | | | | | <0.0001 |
| Normal | 17,935 | 20,977 | 38,912 | 53.9% | 51.1% | |
| Manages to hear | 10,771 | 11,823 | 22,594 | 52.3% | 29.7% | |
| Can hear loud sounds | 6,039 | 7,132 | 13,171 | 54.1% | 17.3% | |
| Poor hearing | 459 | 555 | 1,014 | 54.7% | 1.3% | |

| Variables | Stable or Improved (care needs remained the same or decreased) | Deteriorated (care needs increased) | Total | Deterioration Proportion (%) | % in each category | P value* |
|---|---|---|---|---|---|---|
| | n = 35,289 | n = 40,813 | n = 76,102 | | | |
| Cannot be judged | 85 | 326 | 411 | 79.3% | 0.5% | |
| Expressing intentions | | | | | | <0.0001 |
| Capable | 27,708 | 30,171 | 57,879 | 52.1% | 76.1% | |
| Sometimes capable | 5,722 | 7,852 | 13,574 | 57.8% | 17.8% | |
| Almost incapable | 1,616 | 2,116 | 3,732 | 56.7% | 4.9% | |
| Incapable | 243 | 674 | 917 | 73.5% | 1.2% | |
| Understanding the daily routine | | | | | | <0.0001 |
| Capable | 26,935 | 29,239 | 56,174 | 52.1% | 73.8% | |
| Incapable | 8,354 | 11,574 | 19,928 | 58.1% | 26.2% | |
| Remembering birth date | | | | | | <0.0001 |
| Capable | 32,890 | 36,972 | 69,862 | 52.9% | 91.8% | |
| Incapable | 2,399 | 3,841 | 6,240 | 61.6% | 8.2% | |
| Short-term memory | | | | | | <0.0001 |
| Capable | 23,907 | 26,807 | 50,714 | 52.9% | 66.6% | |
| Incapable | 11,382 | 14,006 | 25,388 | 55.2% | 33.4% | |
| Remembering one’s name | | | | | | <0.0001 |
| Capable | 34,665 | 39,516 | 74,181 | 53.3% | 97.5% | |
| Incapable | 624 | 1,297 | 1,921 | 67.5% | 2.5% | |
| current season | | | | | | <0.0001 |

| Variables | Stable or Improved (care needs remained the same or decreased) | Deteriorated (care needs increased) | Total | Deterioration Proportion (%) | % in each category | P value* |
|---|---|---|---|---|---|---|
| | n = 35,289 | n = 40,813 | n = 76,102 | | | |
| Capable | 29,374 | 32,331 | 61,705 | 52.4% | 81.1% | |
| Incapable | 5,915 | 8,482 | 14,397 | 58.9% | 18.9% | |
| Recognize places | | | | | | <0.0001 |
| Capable | 32,065 | 35,762 | 67,827 | 52.7% | 89.1% | |
| Incapable | 3,224 | 5,051 | 8,275 | 61.0% | 10.9% | |
| Conviction of being threatened | | | | | | <0.0001 |
| Absent | 31,412 | 35,271 | 66,683 | 52.9% | 87.6% | |
| Sometimes present | 1,835 | 2,366 | 4,201 | 56.3% | 5.5% | |
| Present | 2,042 | 3,176 | 5,218 | 60.9% | 6.9% | |
| Delusional thoughts and imaginary conversation | | | | | | <0.0001 |
| Absent | 29,704 | 33,281 | 62,985 | 52.8% | 82.8% | |
| Sometimes present | 2,308 | 2,887 | 5,195 | 55.6% | 6.8% | |
| Present | 3,277 | 4,645 | 7,922 | 58.6% | 10.4% | |
| Emotional instability | | | | | | <0.0001 |
| Absent | 26,978 | 30,829 | 57,807 | 53.3% | 76.0% | |
| Sometimes present | 3,948 | 4,568 | 8,516 | 53.6% | 11.2% | |
| Present | 4,363 | 5,416 | 9,779 | 55.4% | 12.8% | |
| Day–night reversal | | | | | | <0.0001 |
| Absent | 30,455 | 34,828 | 65,283 | 53.3% | 85.8% | |

| Variables | Stable or Improved (care needs remained the same or decreased)<br>n = 35,289 | Deteriorated (care needs increased)<br>n = 40,813 | Total<br>n = 76,102 | Deterioration Proportion (%) | % in each category | P value* |
|---|---|---|---|---|---|---|
| Sometimes present | 2,440 | 2,936 | 5,376 | 54.6% | 7.1% | |
| Present | 2,394 | 3,049 | 5,443 | 56.0% | 7.2% | |
| Speech repetition | | | | | | <0.0001 |
| Absent | 23,960 | 26,976 | 50,936 | 53.0% | 66.9% | |
| Sometimes present | 3,653 | 4,102 | 7,755 | 52.9% | 10.2% | |
| Present | 7,676 | 9,735 | 17,411 | 55.9% | 22.9% | |
| Screaming/Calling out | | | | | | <0.0001 |
| Absent | 31,584 | 35,951 | 67,535 | 53.2% | 88.7% | |
| Sometimes present | 1,894 | 2,383 | 4,277 | 55.7% | 5.6% | |
| Present | 1,811 | 2,479 | 4,290 | 57.8% | 5.6% | |
| Resistance to care | | | | | | <0.0001 |
| Absent | 30,242 | 33,641 | 63,883 | 52.7% | 83.9% | |
| Sometimes present | 2,469 | 3,177 | 5,646 | 56.3% | 7.4% | |
| Present | 2,578 | 3,995 | 6,573 | 60.8% | 8.6% | |
| Frequent wandering | | | | | | <0.0001 |
| Absent | 34,046 | 38,298 | 72,344 | 52.9% | 95.1% | |
| Sometimes present | 479 | 887 | 1,366 | 64.9% | 1.8% | |
| Present | 764 | 1,628 | 2,392 | 68.1% | 3.1% | |
| Restlessness | | | | | | <0.0001 |

| Variables | Stable or Improved (care needs remained the same or decreased) | Deteriorated (care needs increased) | Total | Deterioration Proportion (%) | % in each category | P value* |
|---|---|---|---|---|---|---|
| | n = 35,289 | n = 40,813 | n = 76,102 | | | |
| Absent | 33,514 | 37,996 | 71,510 | 53.1% | 94.0% | |
| Sometimes present | 818 | 1,160 | 1,978 | 58.6% | 2.6% | |
| Present | 957 | 1,657 | 2,614 | 63.4% | 3.4% | |
| Not returning after going out | | | | | | <0.0001 |
| Absent | 34,244 | 38,805 | 73,049 | 53.1% | 96.0% | |
| Sometimes present | 446 | 944 | 1,390 | 67.9% | 1.8% | |
| Present | 599 | 1,064 | 1,663 | 64.0% | 2.2% | |
| Insistence on going out alone | | | | | | <0.0001 |
| Absent | 33,970 | 38,439 | 72,409 | 53.1% | 95.1% | |
| Sometimes present | 625 | 1,035 | 1,660 | 62.3% | 2.2% | |
| Present | 694 | 1,339 | 2,033 | 65.9% | 2.7% | |
| Hoarding behavior | | | | | | <0.0001 |
| Absent | 34,130 | 38,892 | 73,022 | 53.3% | 96.0% | |
| Sometimes present | 350 | 587 | 937 | 62.6% | 1.2% | |
| Present | 809 | 1,334 | 2,143 | 62.2% | 2.8% | |
| Destroying objects and clothing | | | | | | <0.0001 |
| Absent | 34,839 | 40,015 | 74,854 | 53.5% | 98.4% | |
| Sometimes present | 234 | 412 | 646 | 63.8% | 0.8% | |
| Present | 216 | 386 | 602 | 64.1% | 0.8% | |

| Variables | Stable or Improved (care needs remained the same or decreased) | Deteriorated (care needs increased) | Total | Deterioration Proportion (%) | % in each category | P value* |
|---|---|---|---|---|---|---|
| | n = 35,289 | n = 40,813 | n = 76,102 | | | |
| Movement | | | | | | <0.0001 |
| Independent | 17,281 | 23,284 | 40,565 | 57.4% | 53.3% | |
| Needs supervision | 7,892 | 9,782 | 17,674 | 55.3% | 23.2% | |
| Partial assistance | 5,667 | 4,286 | 9,953 | 43.1% | 13.1% | |
| Fully assisted | 4,449 | 3,461 | 7,910 | 43.8% | 10.4% | |
| Passing urine/Urination | | | | | | <0.0001 |
| Independent | 20,617 | 25,710 | 46,327 | 55.5% | 60.9% | |
| Needs supervision | 2,192 | 3,212 | 5,404 | 59.4% | 7.1% | |
| Partial assistance | 7,428 | 7,679 | 15,107 | 50.8% | 19.9% | |
| Fully assisted | 5,052 | 4,212 | 9,264 | 45.5% | 12.2% | |
| Passing motion/Defecation | | | | | | <0.0001 |
| Independent | 21,658 | 27,468 | 49,126 | 55.9% | 64.6% | |
| Needs supervision | 1,975 | 3,029 | 5,004 | 60.5% | 6.6% | |
| Partial assistance | 6,465 | 6,017 | 12,482 | 48.2% | 16.4% | |
| Fully assisted | 5,191 | 4,299 | 9,490 | 45.3% | 12.5% | |
| Everyday decision-making | | | | | | <0.0001 |
| Capable | 12,221 | 13,789 | 26,010 | 53.0% | 34.2% | |
| Capable except during special occasions | 16,855 | 18,017 | 34,872 | 51.7% | 45.8% | |
| Capable but very challenging | 5,314 | 7,423 | 12,737 | 58.3% | 16.7% | |

| Variables | Stable or Improved (care needs remained the same or decreased)<br>n = 35,289 | Deteriorated (care needs increased)<br>n = 40,813 | Total<br>n = 76,102 | Deterioration Proportion (%) | % in each category | P value* |
|---|---|---|---|---|---|---|
| Incapable | 899 | 1,584 | 2,483 | 63.8% | 3.3% | |
| Forgetfulness | | | | | | <0.0001 |
| Absent | 18,446 | 19,587 | 38,033 | 51.5% | 50.0% | |
| Sometimes present | 6,553 | 7,699 | 14,252 | 54.0% | 18.7% | |
| Present | 10,290 | 13,527 | 23,817 | 56.8% | 31.3% | |
| Frequency of going out | | | | | | <0.0001 |
| More than once a week | 19,802 | 22,323 | 42,125 | 53.0% | 55.4% | |
| More than once a month | 6,039 | 7,625 | 13,664 | 55.8% | 18.0% | |
| Less than once a month | 9,448 | 10,865 | 20,313 | 53.5% | 26.7% | |
| Self-talk/Laughing to oneself | | | | | | <0.0001 |
| Absent | 32,938 | 37,637 | 70,575 | 53.3% | 92.7% | |
| Sometimes present | 880 | 1,102 | 1,982 | 55.6% | 2.6% | |
| Present | 1,471 | 2,074 | 3,545 | 58.5% | 4.7% | |
| Selfish behaviors | | | | | | <0.0001 |
| Absent | 31,122 | 35,095 | 66,217 | 53.0% | 87.0% | |
| Sometimes present | 1,918 | 2,379 | 4,297 | 55.4% | 5.6% | |
| Present | 2,249 | 3,339 | 5,588 | 59.8% | 7.3% | |
| Disorganized talking | | | | | | <0.0001 |
| Absent | 27,869 | 31,039 | 58,908 | 52.7% | 77.4% | |

| Variables | Stable or Improved (care needs remained the same or decreased) | Deteriorated (care needs increased) | Total | Deterioration Proportion (%) | % in each category | P value* |
|---|---|---|---|---|---|---|
| | n = 35,289 | n = 40,813 | n = 76,102 | | | |
| Sometimes present | 3,322 | 4,069 | 7,391 | 55.1% | 9.7% | |
| Present | 4,098 | 5,705 | 9,803 | 58.2% | 12.9% | |
| Not fitting in a group | | | | | | <0.0001 |
| Absent | 32,789 | 37,536 | 70,325 | 53.4% | 92.4% | |
| Sometimes present | 1,160 | 1,501 | 2,661 | 56.4% | 3.5% | |
| Present | 1,340 | 1,776 | 3,116 | 57.0% | 4.1% | |
| Shopping | | | | | | <0.0001 |
| Independent | 5,586 | 5,549 | 11,135 | 49.8% | 14.6% | |
| Needs supervision | 369 | 385 | 754 | 51.1% | 1.0% | |
| Partial assistance | 7,777 | 9,245 | 17,022 | 54.3% | 22.4% | |
| Fully assisted | 21,557 | 25,634 | 47,191 | 54.3% | 62.0% | |
| Cooking | | | | | | <0.0001 |
| Independent | 9,788 | 10,502 | 20,290 | 51.8% | 26.7% | |
| Needs supervision | 527 | 679 | 1,206 | 56.3% | 1.6% | |
| Partial assistance | 3,278 | 4,241 | 7,519 | 56.4% | 9.9% | |
| Fully assisted | 21,696 | 25,391 | 47,087 | 53.9% | 61.9% | |
| BPSD_visual/auditory hallucination | | | | | | <0.0001 |
| No | 34,530 | 39,642 | 74,172 | 53.4% | 97.5% | |
| Yes | 759 | 1,171 | 1,930 | 60.7% | 2.5% | |

| Variables | Stable or Improved (care needs remained the same or decreased) | Deteriorated (care needs increased) | Total | Deterioration Proportion (%) | % in each category | P value* |
|---|---|---|---|---|---|---|
| | n = 35,289 | n = 40,813 | n = 76,102 | | | |
| BPSD_ Delusions | | | | | | <0.0001 |
| No | 33,942 | 38,683 | 72,625 | 53.3% | 95.4% | |
| Yes | 1,347 | 2,130 | 3,477 | 61.3% | 4.6% | |
| BPSD_Day-night reversal | | | | | | <0.0001 |
| No | 34,586 | 39,671 | 74,257 | 53.4% | 97.6% | |
| Yes | 703 | 1,142 | 1,845 | 61.9% | 2.4% | |
| BPSD_Violent language | | | | | | <0.0001 |
| No | 34,743 | 39,828 | 74,571 | 53.4% | 98.0% | |
| Yes | 546 | 985 | 1,531 | 64.3% | 2.0% | |
| BPSD_Assaulting | | | | | | <0.0001 |
| No | 35,114 | 40,480 | 75,594 | 53.5% | 99.3% | |
| Yes | 175 | 333 | 508 | 65.6% | 0.7% | |
| BPSD_ Refusal to care | | | | | | <0.0001 |
| No | 34,496 | 39,444 | 73,940 | 53.3% | 97.2% | |
| Yes | 793 | 1,369 | 2,162 | 63.3% | 2.8% | |
| BPSD_Wandering | | | | | | <0.0001 |
| No | 34,823 | 39,891 | 74,714 | 53.4% | 98.2% | |
| Yes | 466 | 922 | 1,388 | 66.4% | 1.8% | |
| BPSD_Mismanagement of fire | | | | | | <0.0001 |

| Variables | Stable or Improved (care needs remained the same or decreased) n = 35,289 | Deteriorated (care needs increased) n = 40,813 | Total n = 76,102 | Deterioration Proportion (%) | % in each category | P value* |
|---|---|---|---|---|---|---|
| No | 34,915 | 39,961 | 74,876 | 53.4% | 98.4% | |
| Yes | 374 | 852 | 1,226 | 69.5% | 1.6% | |
| BPSD_ Filthy behaviors | | | | | | <0.0001 |
| No | 35,028 | 40,359 | 75,387 | 53.5% | 99.1% | |
| Yes | 261 | 454 | 715 | 63.5% | 0.9% | |
| BPSD_Pica behaviors | | | | | | 0.084 |
| No | 35,227 | 40,718 | 75,945 | 53.6% | 99.8% | |
| Yes | 62 | 95 | 157 | 60.5% | 0.2% | |
| BPSD_Sexual disinhibition | | | | | | 0.792 |
| No | 35,274 | 40,794 | 76,068 | 53.6% | 100.0% | |
| Yes | 15 | 19 | 34 | 55.9% | 0.0% | |
| BPSD_Others | | | | | | <0.0001 |
| No | 34,576 | 39,813 | 74,389 | 53.5% | 97.7% | |
| Yes | 713 | 1,000 | 1,713 | 58.4% | 2.3% | |
| No paralysis in limbs | | | | | | <0.0001 |
| No | 24,377 | 26,676 | 51,053 | 52.3% | 67.1% | |
| Yes | 10,912 | 14,137 | 25,049 | 56.4% | 32.9% | |
| Paralysis in left upper limb | | | | | | <0.0001 |
| No | 30,832 | 36,628 | 67,460 | 54.3% | 88.6% | |

| Variables | Stable or Improved (care needs remained the same or decreased) | Deteriorated (care needs increased) | Total | Deterioration Proportion (%) | % in each category | P value* |
|---|---|---|---|---|---|---|
| | n = 35,289 | n = 40,813 | n = 76,102 | | | |
| Yes | 4,457 | 4,185 | 8,642 | 48.4% | 11.4% | |
| Paralysis in right upper limb | | | | | | <0.0001 |
| No | 30,363 | 36,113 | 66,476 | 54.3% | 87.4% | |
| Yes | 4,926 | 4,700 | 9,626 | 48.8% | 12.6% | |
| Paralysis in left lower limb | | | | | | <0.0001 |
| No | 14,736 | 17,590 | 32,326 | 54.4% | 42.5% | |
| Yes | 20,553 | 23,223 | 43,776 | 53.0% | 57.5% | |
| Paralysis in right lower limb | | | | | | 0.014 |
| No | 14,950 | 17,652 | 32,602 | 54.1% | 42.8% | |
| Yes | 20,339 | 23,161 | 43,500 | 53.2% | 57.2% | |
| Paralysis outside limbs | | | | | | <0.0001 |
| No | 31,478 | 37,246 | 68,724 | 54.2% | 90.3% | |
| Yes | 3,811 | 3,567 | 7,378 | 48.3% | 9.7% | |
| Joint movement without restriction | | | | | | <0.0001 |
| No | 19,301 | 19,050 | 38,351 | 49.7% | 50.4% | |
| Yes | 15,988 | 21,763 | 37,751 | 57.6% | 49.6% | |
| Shoulder joint movement restriction | | | | | | <0.0001 |
| No | 29,180 | 35,623 | 64,803 | 55.0% | 85.2% | |
| Yes | 6,109 | 5,190 | 11,299 | 45.9% | 14.8% | |

| Variables | Stable or Improved (care needs remained the same or decreased) | Deteriorated (care needs increased) | Total | Deterioration Proportion (%) | % in each category | P value* |
|---|---|---|---|---|---|---|
| | n = 35,289 | n = 40,813 | n = 76,102 | | | |
| Hip joint movement restriction | | | | | | <0.0001 |
| No | 31,488 | 37,388 | 68,876 | 54.3% | 90.5% | |
| Yes | 3,801 | 3,425 | 7,226 | 47.4% | 9.5% | |
| Knee joint movement restriction | | | | | | <0.0001 |
| No | 24,019 | 29,199 | 53,218 | 54.9% | 69.9% | |
| Yes | 11,270 | 11,614 | 22,884 | 50.8% | 30.1% | |
| Other joint movement restriction | | | | | | <0.0001 |
| No | 26,891 | 32,369 | 59,260 | 54.6% | 77.9% | |
| Yes | 8,398 | 8,444 | 16,842 | 50.1% | 22.1% | |
| Medical treatment in the last 2 weeks_Intravenous therapy | | | | | | <0.0001 |
| No | 34,348 | 40,002 | 74,350 | 53.8% | 97.7% | |
| Yes | 941 | 811 | 1,752 | 46.3% | 2.3% | |
| Medical treatment in the last 2 weeks_Total parenteral nutrition | | | | | | 0.001 |
| No | 35,252 | 40,731 | 75,983 | 53.6% | 99.8% | |
| Yes | 37 | 82 | 119 | 68.9% | 0.2% | |
| Medical treatment in the last 2 weeks_Dialysis | | | | | | <0.0001 |
| No | 34,840 | 40,410 | 75,250 | 53.7% | 98.9% | |
| Yes | 449 | 403 | 852 | 47.3% | 1.1% | |
| Medical treatment in the last 2 weeks_Stoma | | | | | | |

| Variables | Stable or Improved (care needs remained the same or decreased) | Deteriorated (care needs increased) | Total | Deterioration Proportion (%) | % in each category | P value* |
|---|---|---|---|---|---|---|
| | n = 35,289 | n = 40,813 | n = 76,102 | | | |
| No | 35,070 | 40,631 | 75,701 | 53.7% | 99.5% | |
| Yes | 219 | 182 | 401 | 45.4% | 0.5% | |
| Medical treatment in the last 2 weeks_Oxygen therapy | | | | | | 0.001 |
| No | 34,683 | 39,982 | 74,665 | 53.5% | 98.1% | |
| Yes | 606 | 831 | 1,437 | 57.8% | 1.9% | |
| Medical treatment in the last 2 weeks_Artificial respirator | | | | | | 0.345 |
| No | 35,273 | 40,788 | 76,061 | 53.6% | 99.9% | |
| Yes | 16 | 25 | 41 | 61.0% | 0.1% | |
| Medical treatment in the last 2 weeks_Tracheostomy | | | | | | <0.0001 |
| No | 35,262 | 40,745 | 76,007 | 53.6% | 99.9% | |
| Yes | 27 | 68 | 95 | 71.6% | 0.1% | |
| Medical treatment in the last 2 weeks_Pain care | | | | | | 0.009 |
| No | 35,065 | 40,488 | 75,553 | 53.6% | 99.3% | |
| Yes | 224 | 325 | 549 | 59.2% | 0.7% | |
| Medical treatment in the last 2 weeks_Tube feeding | | | | | | <0.0001 |
| No | 35,108 | 40,151 | 75,259 | 53.4% | 98.9% | |
| Yes | 181 | 662 | 843 | 78.5% | 1.1% | |
| Medical treatment in the last 2 weeks_Vital sign monitor | | | | | | <0.0001 |
| No | 35,020 | 40,619 | 75,639 | 53.7% | 99.4% | |

| Variables | Stable or Improved (care needs remained the same or decreased) | Deteriorated (care needs increased) | Total | Deterioration Proportion (%) | % in each category | P value* |
|---|---|---|---|---|---|---|
| | n = 35,289 | n = 40,813 | n = 76,102 | | | |
| Yes | 269 | 194 | 463 | 41.9% | 0.6% | |
| Medical treatment in the last 2 weeks_Bedsore treatment | | | | | | 0.001 |
| No | 34,960 | 40,521 | 75,481 | 53.7% | 99.2% | |
| Yes | 329 | 292 | 621 | 47.0% | 0.8% | |
| Medical treatment in the last 2 weeks_Catheter | | | | | | <0.0001 |
| No | 34,600 | 40,301 | 74,901 | 53.8% | 98.4% | |
| Yes | 689 | 512 | 1,201 | 42.6% | 1.6% | |

* *p* values are two-tailed and are considered to be statistically significant when < 0.005.

Results are after conducting a descriptive analysis of the presence or absence of care needs increases of the total study sample and the sub subgroup. We also conducted a single regression model to identify variables that were statistically significant with *p* value < 0.05 and area under ROC curve ≧ 0.53. None of the variables extracted from the long-term care insurance claim data were statistically significant. We used the selected variables to establish our multivariable binary logistic regression model.

Table A.2. Results of Multivariable Logistic Regression Model of All Care Needs Increases (AUC:0.7143).

| Variables | Odds Ratio (95% Confidence Interval) | P Value |
|---|---|---|
| Sex | | |
| Female | Ref | |
| Male | 1.22 (1.17–1.26) | <0.05 |
| Care needs level of 1st judgment | | |
| Care needs level 1 | Ref | |
| Unqualified | 187 (114–308) | <0.05 |
| Support required Level 1 | 2.78 (2.62–2.95) | <0.05 |
| Support required Level 2 | 1.66 (1.58–1.76) | <0.05 |
| Care needs level 2 | 0.3 (0.28–0.32) | <0.05 |
| Care needs level 3 | 0.11 (0.1–0.13) | <0.05 |
| Care needs level 4 | 0.03 (0.02–0.03) | <0.05 |
| Degree of independent living for elderly with disability | | |
| Ａ２ (rarely goes out and naps several times during daytime） | Ref | |
| Dependent | 0.38 (0.30–0.48) | <0.05 |
| Ｊ１（able to go out using public transportation） | 0.41 (0.38–0.44) | <0.05 |
| Ｊ２（able to go out but only within the neighborhood） | 0.57 (0.54–0.60) | <0.05 |
| Ａ１（goes out with assistance and usually not bedridden） | 0.84 (0.81–0.88) | <0.05 |
| Ｂ１（able to eat and use toilet away from bed using wheel chair） | 1.14 (1.06–1.21) | <0.05 |
| Ｂ２（requires assistance to get onto a wheelchair） | 1.17 (1.07–1.29) | <0.05 |
| Ｃ１（able to roll over by him/herself） | 1.66 (1.43–1.93) | <0.05 |
| Ｃ２（fully assisted in rolling over by him/herself） | 1.75 (1.45–2.12) | <0.05 |
| Cognitive ability (daily decision-making) | | |
| Loss | Ref | |
| Unknown | 0.30 (0.04–2.50) | 0.268 |
| Independent | 1.07 (1.03–1.11) | <0.05 |
| Some difficulty | 1.36 (1.28–1.45) | <0.05 |
| Partially assisted | 1.49 (1.36–1.63) | <0.05 |
| Fully assisted | 1.48 (1.29–1.70) | <0.05 |
| Communication ability | | |
| Capable | Ref | |
| Unknown | 1.16 (0.18–7.65) | 0.878 |

| Variables | Odds Ratio (95% Confidence Interval) | P Value |
|---|---|---|
| Somewhat challenging | 0.99 (0.93–1.06) | 0.743 |
| Limited to specific requirements | 0.99 (0.89–1.09) | 0.792 |
| Incapable | 1.03 (0.86–1.23) | 0.747 |
| Turn in bed | | |
| Can do holding railings | Ref | |
| Independent | 1.11 (1.07–1.16) | <0.05 |
| Fully assisted | 1.10 (1.02–1.18) | <0.05 |
| Get up | | |
| Can do holding railings | Ref | |
| Independent | 1.06 (1.00–1.13) | 0.052 |
| Fully assisted | 1.36 (1.25–1.47) | <0.05 |
| Stand on both feet | | |
| Independent | Ref | |
| Partially assisted | 1.11 (1.06–1.16) | <0.05 |
| Fully assisted | 1.11 (1.00–1.25) | 0.057 |
| Walk | | |
| Can do with assistive technology | Ref | |
| Independent | 0.87 (0.83–0.92) | <0.05 |
| Fully assisted | 1.08 (1.01–1.16) | <0.05 |
| Transfer | | |
| Independent | Ref | |
| Needs supervision | 0.97 (0.92–1.03) | 0.328 |
| Partially assisted | 1.04 (0.96–1.13) | 0.367 |
| Fully assisted | 0.98 (0.85–1.14) | 0.835 |
| Stand up | | |
| Can do holding railings | Ref | |
| Independent | 1.39 (1.26–1.53) | <0.05 |
| Fully assisted | 1.14 (1.03–1.26) | <0.05 |
| Stand on one foot | | |
| Partially assisted | Ref | |
| Independent | 1.03 (0.95–1.11) | 0.505 |
| Fully assisted | 1.06 (1.01–1.11) | <0.05 |
| Swallowing | | |

| Variables | Odds Ratio (95% Confidence Interval) | P Value |
|---|---|---|
| Independent | Ref | |
| Needs supervision | 0.96 (0.92–1.00) | <0.05 |
| Fully assisted | 3.10 (2.48–3.88) | <0.05 |
| Washing face | | |
| Independent | Ref | |
| Partially assisted | 1.11 (1.06–1.18) | <0.05 |
| Fully assisted | 1.32 (1.19–1.46) | <0.05 |
| Putting on/removing top | | |
| Independent | Ref | |
| Needs supervision | 1.30 (1.19–1.43) | <0.05 |
| Partially assisted | 1.44 (1.33–1.55) | <0.05 |
| Fully assisted | 1.65 (1.46–1.87) | <0.05 |
| Putting on/removing pants | | |
| Independent | Ref | |
| Needs supervision | 0.97 (0.88–1.06) | 0.487 |
| Partially assisted | 0.99 (0.91–1.07) | 0.812 |
| Fully assisted | 1.08 (0.96–1.22) | 0.21 |
| Expressing intentions | | |
| Capable | Ref | |
| Sometimes capable | 1.26 (1.19–1.32) | <0.05 |
| Almost incapable | 1.40 (1.27–1.54) | <0.05 |
| Incapable | 1.78 (1.45–2.18) | <0.05 |
| Understanding the daily routine | | |
| Capable | Ref | |
| Incapable | 1.29 (1.22–1.36) | <0.05 |
| Remembering birth date | | |
| Capable | Ref | |
| Incapable | 1.18 (1.11–1.25) | <0.05 |
| Recognize places | | |
| Capable | Ref | |
| Incapable | 1.21 (1.12–1.30) | <0.05 |
| Conviction of being threatened | | |
| Absent | Ref | |

| Variables | Odds Ratio (95% Confidence Interval) | P Value |
|---|---|---|
| Sometimes absent | 1.27 (1.19–1.35) | <0.05 |
| Present | 1.45 (1.36–1.54) | <0.05 |
| Movement | | |
| Independent | Ref | |
| Needs supervision | 1.13 (1.07–1.19) | <0.05 |
| Partial assistance | 1.27 (1.18–1.37) | <0.05 |
| Fully assisted | 1.15 (1.03–1.28) | <0.05 |
| Pass motion/Defecation | | |
| Independent | Ref | |
| Needs supervision | 1.32 (1.23–1.42) | <0.05 |
| Partial assistance | 1.64 (1.53–1.76) | <0.05 |
| Fully assisted | 1.78 (1.60–1.97) | <0.05 |
| Disorganized talking | | |
| Absent | Ref | |
| Sometimes absent | 1.25 (1.19–1.32) | <0.05 |
| Present | 1.25 (1.18–1.32) | <0.05 |
| No paralysis in limbs | | |
| No | Ref | |
| Yes | 0.85 (0.81–0.88) | <0.05 |
| Joint movement without restriction | | |
| No | Ref | |
| Yes | 1.19 (1.15–1.23) | <0.05 |
| Shoulder joint movement restriction | | |
| No | Ref | |
| Yes | 0.98 (0.94–1.03) | 0.541 |
| Care needs certification reference time_Dementia addition (minute) | 1.01 (1.01–1.01) | <0.05 |
