## Appendix B Outcomes of the machine learning models for "Development and validation of gradient boosting decision tree models for predicting care needs using a long-term care database in Japan"

Outcomes of the machine learning models with four care needs level (CNL) categories (declined needs and benefit loss as one group, support required levels 1 and 2 into one group, CNL 1–3 as one group, and CNL 4–5 to another group) and nine CNL categories (the original CNL, benefit loss, and declined needs).

Table B.1. Labels of Classification Models.

| # of labels | Label description |
|---|---|
| 4 | 0: Declined needs; Loss of benefits |
| | 1: Support required level 1–2 |
| | 2: Care needs level 1–3 |
| | 3: Care needs level 4–5 |
| 9 | 00: Declined needs |
| | 10: Loss of benefits |
| | 12: Support required level 1 |
| | 13: Support required level 2 |
| | 21: Care needs level 1 |
| | 22: Care needs level 2 |
| | 23: Care needs level 3 |
| | 24: Care needs level 4 |
| | 25: Care needs level 5 |

Table B.2. Demographic Characteristics and Chi-square or Fisher's Exact Test Result of Numbers of Stable or Improved and Deteriorated Groups.

| **Features** | **Level** | **Missing** | **Stable or Improved (care needs remained the same or decreased) n = 18,659** | **Deteriorated (care needs increased) n = 11,728** | **Total N = 30,387** | **Deterioration Proportion (%)** | **% in each category** | **P value** |
|---|---|---|---|---|---|---|---|---|
| Living arrangement | Unknown | 0 | 1 | 0 | 1 | 0.00% | 0.00% | <0.001 |
| | Home | | 17,785 | 11,370 | 29,155 | 39.00% | 95.90% | |
| | facility | | 873 | 358 | 1,231 | 29.10% | 4.10% | |
| Signs of elderly people with dementia | None | 0 | 18,659 | 11,728 | 30,387 | 38.60% | 100.00% | 1 |
| Applicant type | 1st insured | 0 | 18,658 | 11,728 | 30,386 | 38.60% | 100.00% | 1 |
| | 2nd insured | | 1 | | 1 | 0.00% | 0.00% | |
| Sex | Male | 0 | 5,993 | 2,517 | 8,510 | 29.60% | 28.00% | <0.001 |
| | Female | | 12,666 | 9,211 | 21,877 | 42.10% | 72.00% | |
| Care needs level_1st judgment | Unqualified | 0 | 223 | 207 | 430 | 48.10% | 1.40% | <0.001 |
| | Support required level 1 | | 2,315 | 2,106 | 4,421 | 47.60% | 14.50% | |
| | Support required level 2 | | 2,464 | 2,487 | 4,951 | 50.20% | 16.30% | |
| | Care needs level 1 | | 2,973 | 2,459 | 5,432 | 45.30% | 17.90% | |
| | Care needs level 2 | | 3,137 | 2036 | 5,173 | 39.40% | 17.00% | |
| | Care needs level 3 | | 2025 | 1,071 | 3,096 | 34.60% | 10.20% | |
| | Care needs level 4 | | 2,722 | 890 | 3,612 | 24.60% | 11.90% | |
| | Care needs level 5 | | 2,800 | 472 | 3,272 | 14.40% | 10.80% | |

| Features | Level | Missing | Stable or Improved (care needs remained the same or decreased) n = 18,659 | Deteriorated (care needs increased) n = 11,728 | Total N = 30,387 | Deterioration Proportion (%) | % in each category | P value |
|---|---|---|---|---|---|---|---|---|
| Care needs level_2nd judgment | Unqualified | 0 | 26 | 35 | 61 | 57.40% | 0.20% | <0.001 |
| | Support required level 1 | | 2046 | 1,892 | 3,938 | 48.00% | 13.00% | |
| | Support required level 2 | | 2,115 | 2,131 | 4,246 | 50.20% | 14.00% | |
| | Care needs level 1 | | 2,921 | 2,628 | 5,549 | 47.40% | 18.30% | |
| | Care needs level 2 | | 3,068 | 2,116 | 5,184 | 40.80% | 17.10% | |
| | Care needs level 3 | | 2,552 | 1,454 | 4,006 | 36.30% | 13.20% | |
| | Care needs level 4 | | 2,722 | 977 | 3,699 | 26.40% | 12.20% | |
| | Care needs level 5 | | 3,209 | 495 | 3,704 | 13.40% | 12.20% | |
| Year of certification criteria adopted | 2009 | 0 | 8,172 | 3,565 | 11,737 | 30.40% | 38.60% | <0.001 |
| | 2012 | | 10,487 | 8,163 | 18,650 | 43.80% | 61.40% | |
| Turn in bed | Independent | 0 | 5,158 | 4,071 | 9,229 | 44.10% | 30.40% | <0.001 |
| | Can do holding railings | | 9,604 | 6,609 | 16,213 | 40.80% | 53.40% | |
| | Fully assisted | | 3,897 | 1,048 | 4,945 | 21.20% | 16.30% | |
| Get up | Independent | 0 | 1,233 | 947 | 2,180 | 43.40% | 7.20% | <0.001 |
| | Can do holding railings | | 12,765 | 9,659 | 22,424 | 43.10% | 73.80% | |
| | Fully assisted | | 4,661 | 1,122 | 5,783 | 19.40% | 19.00% | |
| Maintain sitting position | Independent | 0 | 5,270 | 4,387 | 9,657 | 45.40% | 31.80% | <0.001 |
| | Can do holding railings | | 6,457 | 4,756 | 11,213 | 42.40% | 36.90% | |

| Features | Level | Missing | Stable or Improved (care needs remained the same or decreased) n = 18,659 | Deteriorated (care needs increased) n = 11,728 | Total N = 30,387 | Deterioration Proportion (%) | % in each category | P value |
|---|---|---|---|---|---|---|---|---|
| | Partially assisted | | 6,024 | 2,470 | 8,494 | 29.10% | 28.00% | |
| | Fully assisted | | 908 | 115 | 1,023 | 11.20% | 3.40% | |
| Sitting, with feet dangling | Independent | 0 | 18,659 | 11,728 | 30,387 | 38.60% | 100.00% | 1 |
| Stand on both feet | Independent | 0 | 6,378 | 5,288 | 11,666 | 45.30% | 38.40% | <0.001 |
| | Partially assisted | | 8,025 | 5,464 | 13,489 | 40.50% | 44.40% | |
| | Fully assisted | | 4,256 | 976 | 5,232 | 18.70% | 17.20% | |
| Walk | Independent | 0 | 3,409 | 2,614 | 6,023 | 43.40% | 19.80% | <0.001 |
| | Can do with assistive technology | | 8,997 | 7,233 | 16,230 | 44.60% | 53.40% | |
| | Fully assisted | | 6,253 | 1,881 | 8,134 | 23.10% | 26.80% | |
| Transfer | Independent | 0 | 8,722 | 7,480 | 16,202 | 46.20% | 53.30% | <0.001 |
| | Needs supervision | | 3,177 | 2,099 | 5,276 | 39.80% | 17.40% | |
| | Partially assisted | | 2,702 | 1,278 | 3,980 | 32.10% | 13.10% | |
| | Fully assisted | | 4,058 | 871 | 4,929 | 17.70% | 16.20% | |
| Stand up | Independent | 0 | 485 | 400 | 885 | 45.20% | 2.90% | <0.001 |
| | Can do holding railings | | 13,465 | 10,181 | 23,646 | 43.10% | 77.80% | |
| | Fully assisted | | 4,709 | 1,147 | 5,856 | 19.60% | 19.30% | |
| Stand on one foot | Independent | 0 | 613 | 459 | 1,072 | 42.80% | 3.50% | <0.001 |
| | Can do holding railings | | 9,228 | 7,502 | 16,730 | 44.80% | 55.10% | |

| **Features** | **Level** | **Missing** | **Stable or Improved (care needs remained the same or decreased) n = 18,659** | **Deteriorated (care needs increased) n = 11,728** | **Total N = 30,387** | **Deterioration Proportion (%)** | **% in each category** | **P value** |
|---|---|---|---|---|---|---|---|---|
| | Fully assisted | | 8,818 | 3,767 | 12,585 | 29.90% | 41.40% | |
| Stepping into a bathtub | Independent | 0 | 18,659 | 11,728 | 30,387 | 38.60% | 100.00% | 1 |
| Bathing | Independent | 0 | 5,112 | 4,728 | 9,840 | 48.00% | 32.40% | <0.001 |
| | Partially assisted | | 5,393 | 4,119 | 9,512 | 43.30% | 31.30% | |
| | Fully assisted | | 6,973 | 2,499 | 9,472 | 26.40% | 31.20% | |
| | Does not take bath | | 1,181 | 382 | 1,563 | 24.40% | 5.10% | |
| Bedsores | Present | 0 | 18,659 | 11,728 | 30,387 | 38.60% | 100.00% | 1 |
| Skin disease | Present | 0 | 18,659 | 11,728 | 30,387 | 38.60% | 100.00% | 1 |
| Raising an arm till chest level | Capable | 0 | 18,659 | 11,728 | 30,387 | 38.60% | 100.00% | 1 |
| Swallowing | Independent | 0 | 11,863 | 8,887 | 20,750 | 42.80% | 68.30% | <0.001 |
| | Needs supervision | | 5,982 | 2,760 | 8,742 | 31.60% | 28.80% | |
| | Fully assisted | | 814 | 81 | 895 | 9.10% | 2.90% | |
| Urination urge | Present | 0 | 18,659 | 11,728 | 30,387 | 38.60% | 100.00% | 1 |
| Bowel movement urge | Present | 0 | 18,659 | 11,728 | 30,387 | 38.60% | 100.00% | 1 |
| Cleaning after urination | Independent | 0 | 18,659 | 11,728 | 30,387 | 38.60% | 100.00% | 1 |
| Cleaning after bowel movement | Independent | 0 | 18,659 | 11,728 | 30,387 | 38.60% | 100.00% | 1 |
| Eating | Independent | 0 | 12,166 | 9,628 | 21,794 | 44.20% | 71.70% | <0.001 |
| | Needs supervision | | 2,395 | 1,195 | 3,590 | 33.30% | 11.80% | |

| Features | Level | Missing | Stable or Improved (care needs remained the same or decreased) n = 18,659 | Deteriorated (care needs increased) n = 11,728 | Total N = 30,387 | Deterioration Proportion (%) | % in each category | P value |
|---|---|---|---|---|---|---|---|---|
| | Partially assisted | | 1,791 | 584 | 2,375 | 24.60% | 7.80% | |
| | Fully assisted | | 2,307 | 321 | 2,628 | 12.20% | 8.60% | |
| Oral hygiene | Independent | 0 | 9,663 | 8,356 | 18,019 | 46.40% | 59.30% | <0.001 |
| | Partially assisted | | 4,925 | 2,468 | 7,393 | 33.40% | 24.30% | |
| | Fully assisted | | 4,071 | 904 | 4,975 | 18.20% | 16.40% | |
| Washing face | Independent | 0 | 9,895 | 8,481 | 18,376 | 46.20% | 60.50% | <0.001 |
| | Partially assisted | | 4,733 | 2,371 | 7,104 | 33.40% | 23.40% | |
| | Fully assisted | | 4,031 | 876 | 4,907 | 17.90% | 16.10% | |
| Hair care | Independent | 0 | 10,727 | 8,885 | 19,612 | 45.30% | 64.50% | <0.001 |
| | Partially assisted | | 3,122 | 1,606 | 4,728 | 34.00% | 15.60% | |
| | Fully assisted | | 4,810 | 1,237 | 6,047 | 20.50% | 19.90% | |
| Cutting nails | Independent | 0 | 5,116 | 4,543 | 9,659 | 47.00% | 31.80% | <0.001 |
| | Partially assisted | | 3,683 | 2,912 | 6,595 | 44.20% | 21.70% | |
| | Fully assisted | | 9,860 | 4,273 | 14,133 | 30.20% | 46.50% | |
| Buttoning/unbuttoning clothes | Unknown | 0 | 18,659 | 11,728 | 30,387 | 38.60% | 100.00% | 1 |
| Putting on/ removing top | Independent | 0 | 8,022 | 7,168 | 15,190 | 47.20% | 50.00% | <0.001 |
| | Needs supervision | | 1,670 | 1,239 | 2,909 | 42.60% | 9.60% | |
| | Partially assisted | | 4,532 | 2,368 | 6,900 | 34.30% | 22.70% | |

| **Features** | **Level** | **Missing** | **Stable or Improved (care needs remained the same or decreased) n = 18,659** | **Deteriorated (care needs increased) n = 11,728** | **Total N = 30,387** | **Deterioration Proportion (%)** | **% in each category** | **P value** |
|---|---|---|---|---|---|---|---|---|
| | Fully assisted | | 4,435 | 953 | 5,388 | 17.70% | 17.70% | |
| Putting on/removing pants | Independent | 0 | 7,672 | 6,916 | 14,588 | 47.40% | 48.00% | <0.001 |
| | Needs supervision | | 1,575 | 1,203 | 2,778 | 43.30% | 9.10% | |
| | Partially assisted | | 3,851 | 2,228 | 6,079 | 36.70% | 20.00% | |
| | Fully assisted | | 5,561 | 1,381 | 6,942 | 19.90% | 22.80% | |
| Putting on/removing socks | Unknown | 0 | 18,659 | 11,728 | 30,387 | 38.60% | 100.00% | 1 |
| Cleaning up rooms | Unknown | 0 | 18,659 | 11,728 | 30,387 | 38.60% | 100.00% | 1 |
| Taking medication | Independent | 0 | 5,798 | 5,138 | 10,936 | 47.00% | | <0.001 |
| | Partially assisted | | 7,106 | 4,847 | 11,953 | 40.60% | 39.30% | |
| | Fully assisted | | 5,755 | 1,743 | 7,498 | 23.20% | 24.70% | |
| Managing money | Independent | 0 | 4,699 | 4,264 | 8,963 | 47.60% | 29.50% | <0.001 |
| | Partially assisted | | 3,088 | 2,489 | 5,577 | 44.60% | 18.40% | |
| | Fully assisted | | 10,872 | 4,975 | 15,847 | 31.40% | 52.20% | |
| Severe forgetfulness | Absent | 0 | 18,659 | 11,728 | 30,387 | 38.60% | 100.00% | 1 |
| Indifference to surroundings | Absent | 0 | 18,659 | 11,728 | 30,387 | 38.60% | 100.00% | 1 |
| Vision | Normal | 0 | 11,021 | 7,397 | 18,418 | 40.20% | 60.60% | <0.001 |
| | Able to see over 1m away | | 5,294 | 3,403 | 8,697 | 39.10% | 28.60% | |
| | Nearsightedness | | 1,270 | 692 | 1962 | 35.30% | 6.50% | |

| Features | Level | Missing | Stable or Improved (care needs remained the same or decreased) n = 18,659 | Deteriorated (care needs increased) n = 11,728 | Total N = 30,387 | Deterioration Proportion (%) | % in each category | P value |
|---|---|---|---|---|---|---|---|---|
| | Poor sight | | 368 | 144 | 512 | 28.10% | 1.70% | |
| | Cannot be judged | | 706 | 92 | 798 | 11.50% | 2.60% | |
| Hearing ability | Normal | 0 | 7,600 | 5,064 | 12,664 | 40.00% | 41.70% | <0.001 |
| | Manages to hear | | 5,849 | 3,811 | 9,660 | 39.50% | 31.80% | |
| | Able to hear loud sounds | | 4,359 | 2,614 | 6,973 | 37.50% | 22.90% | |
| | Poor hearing | | 436 | 195 | 631 | 30.90% | 2.10% | |
| | Cannot be judged | | 415 | 44 | 459 | 9.60% | 1.50% | |
| Expressing intentions | Capable | 0 | 11,419 | 8,855 | 20,274 | 43.70% | 66.70% | <0.001 |
| | Sometimes capable | | 4,066 | 2033 | 6,099 | 33.30% | 20.10% | |
| | Almost incapable | | 2,078 | 684 | 2,762 | 24.80% | 9.10% | |
| | Incapable | | 1,096 | 156 | 1,252 | 12.50% | 4.10% | |
| Responding to request | Capable | 0 | 18,659 | 11,728 | 30,387 | 38.60% | 100.00% | 1 |
| Understanding the daily routine | Capable | 0 | 10,814 | 8,507 | 19,321 | 44.00% | 63.60% | <0.001 |
| | Incapable | | 7,845 | 3,221 | 11,066 | 29.10% | 36.40% | |
| Remembering birth date | Capable | 0 | 15,018 | 10,641 | 25,659 | 41.50% | 84.40% | <0.001 |
| | Incapable | | 3,641 | 1,087 | 4,728 | 23.00% | 15.60% | |
| Short-term memory | Capable | 0 | 9,929 | 7,622 | 17,551 | 43.40% | 57.80% | <0.001 |
| | Incapable | | 8,730 | 4,106 | 12,836 | 32.00% | 42.20% | |

| **Features** | **Level** | **Missing** | **Stable or Improved (care needs remained the same or decreased) n = 18,659** | **Deteriorated (care needs increased) n = 11,728** | **Total N = 30,387** | **Deterioration Proportion (%)** | **% in each category** | **P value** |
|---|---|---|---|---|---|---|---|---|
| Remembering one's name | Capable | 0 | 16,886 | 11,380 | 28,266 | 40.30% | 93.00% | <0.001 |
| | Incapable | | 1,773 | 348 | 2,121 | 16.40% | 7.00% | |
| Aware of current season | Capable | 0 | 12,475 | 9,337 | 21,812 | 42.80% | 71.80% | <0.001 |
| | Incapable | | 6,184 | 2,391 | 8,575 | 27.90% | 28.20% | |
| Recognizing places | Capable | 0 | 14,254 | 10,310 | 24,564 | 42.00% | 80.80% | <0.001 |
| | Incapable | | 4,405 | 1,418 | 5,823 | 24.40% | 19.20% | |
| Conviction to be threatened | Absent | 0 | 16,313 | 10,135 | 26,448 | 38.30% | 87.00% | 0.005 |
| | Sometimes present | | 1,026 | 646 | 1,672 | 38.60% | 5.50% | |
| | Present | | 1,320 | 947 | 2,267 | 41.80% | 7.50% | |
| Delusional thoughts and imaginary conversation | Absent | 0 | 15,327 | 9,578 | 24,905 | 38.50% | 82.00% | 0.137 |
| | Sometimes present | | 1,286 | 879 | 2,165 | 40.60% | 7.10% | |
| | Present | | 2046 | 1,271 | 3,317 | 38.30% | 10.90% | |
| Illusion | Absent | 0 | 18,659 | 11,728 | 30,387 | 38.60% | 100.00% | 1 |
| Emotional instability | Absent | 0 | 14,075 | 8,873 | 22,948 | 38.70% | 75.50% | 0.004 |
| | Sometimes present | | 2017 | 1,367 | 3,384 | 40.40% | 11.10% | |
| | Present | | 2,567 | 1,488 | 4,055 | 36.70% | 13.30% | |
| Day–night reversal | Absent | 0 | 15,084 | 9,970 | 25,054 | 39.80% | 82.40% | <0.001 |
| | Sometimes present | | 1,680 | 904 | 2,584 | 35.00% | 8.50% | |

| Features | Level | Missing | Stable or Improved (care needs remained the same or decreased) n = 18,659 | Deteriorated (care needs increased) n = 11,728 | Total N = 30,387 | Deterioration Proportion (%) | % in each category | P value |
|---|---|---|---|---|---|---|---|---|
| | Present | | 1,895 | 854 | 2,749 | 31.10% | 9.00% | |
| Violent behaviors | Unknown | 0 | 18,659 | 11,728 | 30,387 | 38.60% | 100.00% | 1 |
| Speech repetition | Absent | 0 | 12,311 | 7,297 | 19,608 | 37.20% | 64.50% | <0.001 |
| | Sometimes present | | 1922 | 1,401 | 3,323 | 42.20% | 10.90% | |
| | Present | | 4,426 | 3,030 | 7,456 | 40.60% | 24.50% | |
| Screaming/Calling out | Absent | 0 | 15,957 | 10,412 | 26,369 | 39.50% | 86.80% | <0.001 |
| | Sometimes present | | 1,274 | 642 | 1916 | 33.50% | 6.30% | |
| | Present | | 1,428 | 674 | 2,102 | 32.10% | 6.90% | |
| Resistance to care | Absent | 0 | 14,698 | 9,702 | 24,400 | 39.80% | 80.30% | <0.001 |
| | Sometimes present | | 1,675 | 957 | 2,632 | 36.40% | 8.70% | |
| | Present | | 2,286 | 1,069 | 3,355 | 31.90% | 11.00% | |
| Frequent wandering | Absent | 0 | 17,567 | 11,161 | 28,728 | 38.90% | 94.50% | <0.001 |
| | Sometimes present | | 353 | 223 | 576 | 38.70% | 1.90% | |
| | Present | | 739 | 344 | 1,083 | 31.80% | 3.60% | |
| Restlessness | Absent | 0 | 17,224 | 10,936 | 28,160 | 38.80% | 92.70% | 0.007 |
| | Sometimes present | | 620 | 327 | 947 | 34.50% | 3.10% | |
| | Present | | 815 | 465 | 1,280 | 36.30% | 4.20% | |
| Not returning after going out | Absent | 0 | 17,889 | 11,234 | 29,123 | 38.60% | 95.80% | 0.238 |

| Features | Level | Missing | Stable or Improved (care needs remained the same or decreased) n = 18,659 | Deteriorated (care needs increased) n = 11,728 | Total N = 30,387 | Deterioration Proportion (%) | % in each category | P value |
|---|---|---|---|---|---|---|---|---|
| | Sometimes absent | | 293 | 211 | 504 | 41.90% | 1.70% | |
| | Present | | 477 | 283 | 760 | 37.20% | 2.50% | |
| Insistence on going out alone | Absent | 0 | 17,642 | 11,154 | 28,796 | 38.70% | 94.80% | 0.011 |
| | Sometimes present | | 429 | 274 | 703 | 39.00% | 2.30% | |
| | Present | | 588 | 300 | 888 | 33.80% | 2.90% | |
| Hoarding behavior | Absent | 0 | 17,903 | 11,211 | 29,114 | 38.50% | 95.80% | 0.108 |
| | Sometimes present | | 244 | 147 | 391 | 37.60% | 1.30% | |
| | Present | | 512 | 370 | 882 | 42.00% | 2.90% | |
| Mismanagement of fire | Unknown | 0 | 18,659 | 11,728 | 30,387 | 38.60% | 100.00% | 1 |
| Destroying objects and clothing | Absent | 0 | 18,305 | 11,528 | 29,833 | 38.60% | 98.20% | 0.3 |
| | Sometimes present | | 160 | 99 | 259 | 38.20% | 0.90% | |
| | Present | | 194 | 101 | 295 | 34.20% | 1.00% | |
| Unhygienic behaviors | Unknown | 0 | 18,659 | 11,728 | 30,387 | 38.60% | 100.00% | 1 |
| Pica behaviors | Unknown | 0 | 18,659 | 11,728 | 30,387 | 38.60% | 100.00% | 1 |
| Inappropriate sexual behavior | Unknown | 0 | 18,659 | 11,728 | 30,387 | 38.60% | 100.00% | 1 |
| Movement | Independent | 0 | 7,097 | 6,243 | 13,340 | 46.80% | 43.90% | <0.001 |
| | Needs supervision | | 3,832 | 2,805 | 6,637 | 42.30% | 21.80% | |
| | Partially assisted | | 2,814 | 1,456 | 4,270 | 34.10% | 14.10% | |

| Features | Level | Missing | Stable or Improved (care needs remained the same or decreased) n = 18,659 | Deteriorated (care needs increased) n = 11,728 | Total N = 30,387 | Deterioration Proportion (%) | % in each category | P value |
|---|---|---|---|---|---|---|---|---|
| | Fully assisted | | 4,916 | 1,224 | 6,140 | 19.90% | 20.20% | |
| Cleaning after urination | Unknown | 0 | 18,659 | 11,728 | 30,387 | 38.60% | 100.00% | 1 |
| Passing urine/ Urination | Independent | 0 | 8,030 | 7,216 | 15,246 | 47.30% | 50.20% | <0.001 |
| | Needs supervision | | 1,225 | 880 | 2,105 | 41.80% | 6.90% | |
| | Partially assisted | | 3,766 | 2,167 | 5,933 | 36.50% | 19.50% | |
| | Fully assisted | | 5,638 | 1,465 | 7,103 | 20.60% | 23.40% | |
| Passing motion/ Defecation | Independent | 0 | 8,606 | 7,582 | 16,188 | 46.80% | 53.30% | <0.001 |
| | Needs supervision | | 1,187 | 807 | 1994 | 40.50% | 6.60% | |
| | Partially assisted | | 3,190 | 1,858 | 5,048 | 36.80% | 16.60% | |
| | Fully assisted | | 5,676 | 1,481 | 7,157 | 20.70% | 23.60% | |
| Managing money | Unknown | 0 | 18,659 | 11,728 | 30,387 | 38.60% | 100.00% | 1 |
| Everyday decision-making | Capable | 0 | 4,761 | 3,976 | 8,737 | 45.50% | 28.80% | <0.001 |
| | Capable except during special occasions | | 7,322 | 5,246 | 12,568 | 41.70% | 41.40% | |
| | Capable but very challenging | | 4,396 | 2045 | 6,441 | 31.70% | 21.20% | |
| | Incapable | | 2,180 | 461 | 2,641 | 17.50% | 8.70% | |
| Forgetfulness | Absent | 0 | 8,570 | 5,198 | 13,768 | 37.80% | 45.30% | <0.001 |
| | Sometimes present | | 3,234 | 2,464 | 5,698 | 43.20% | 18.80% | |
| | Present | | 6,855 | 4,066 | 10,921 | 37.20% | 35.90% | |

| Features | Level | Missing | Stable or Improved (care needs remained the same or decreased) n = 18,659 | Deteriorated (care needs increased) n = 11,728 | Total N = 30,387 | Deterioration Proportion (%) | % in each category | P value |
|---|---|---|---|---|---|---|---|---|
| Activity level during daytime | Unknown | 0 | 18,659 | 11,728 | 30,387 | 38.60% | 100.00% | 1 |
| Frequency of going out | More than once a week | 0 | 8,683 | 7,055 | 15,738 | 44.80% | 51.80% | <0.001 |
| | More than once a month | | 2,811 | 1972 | 4,783 | 41.20% | 15.70% | |
| | Less than once a month | | 7,165 | 2,701 | 9,866 | 27.40% | 32.50% | |
| Change in environment | Unknown | 0 | 18,659 | 11,728 | 30,387 | 38.60% | 100.00% | 1 |
| Self-talk / laughing to oneself | Absent | 0 | 16,720 | 10,746 | 27,466 | 39.10% | 90.40% | <0.001 |
| | Sometimes present | | 643 | 346 | 989 | 35.00% | 3.30% | |
| | Present | | 1,296 | 636 | 1932 | 32.90% | 6.40% | |
| Selfish behaviors | Absent | 0 | 16,186 | 10,195 | 26,381 | 38.60% | 86.80% | 0.135 |
| | Sometimes present | | 1,021 | 681 | 1,702 | 40.00% | 5.60% | |
| | Present | | 1,452 | 852 | 2,304 | 37.00% | 7.60% | |
| Disorganized talking | Absent | 0 | 13,812 | 8,911 | 22,723 | 39.20% | 74.80% | <0.001 |
| | Sometimes present | | 1,866 | 1,244 | 3,110 | 40.00% | 10.20% | |
| | Present | | 2,981 | 1,573 | 4,554 | 34.50% | 15.00% | |
| Not fitting in a group | Absent | 0 | 16,986 | 10,695 | 27,681 | 38.60% | 91.10% | 0.194 |
| | Sometimes present | | 731 | 487 | 1,218 | 40.00% | 4.00% | |
| | Present | | 942 | 546 | 1,488 | 36.70% | 4.90% | |
| Shopping | Independent | 0 | 1942 | 1,790 | 3,732 | 48.00% | 12.30% | <0.001 |

| **Features** | **Level** | **Missing** | **Stable or Improved (care needs remained the same or decreased) n = 18,659** | **Deteriorated (care needs increased) n = 11,728** | **Total N = 30,387** | **Deterioration Proportion (%)** | **% in each category** | **P value** |
|---|---|---|---|---|---|---|---|---|
| | Needs supervision | | 134 | 149 | 283 | 52.70% | 0.90% | |
| | Partially assisted | | 2,793 | 2,665 | 5,458 | 48.80% | 18.00% | |
| | Fully assisted | | 13,790 | 7,124 | 20,914 | 34.10% | 68.80% | |
| Light cooking | Independent | 0 | 3,624 | 3,446 | 7,070 | 48.70% | 23.30% | <0.001 |
| | Needs supervision | | 199 | 208 | 407 | 51.10% | 1.30% | |
| | Partially assisted | | 1,369 | 1,347 | 2,716 | 49.60% | 8.90% | |
| | Fully assisted | | 13,467 | 6,727 | 20,194 | 33.30% | 66.50% | |
| Bedridden (levels) | Independent | 0 | 53 | 48 | 101 | 47.50% | 0.30% | <0.001 |
| | J1 | | 1,251 | 1,130 | 2,381 | 47.50% | 7.80% | |
| | J2 | | 2,641 | 2,368 | 5,009 | 47.30% | 16.50% | |
| | A1 | | 3,268 | 3,049 | 6,317 | 48.30% | 20.80% | |
| | A2 | | 4,283 | 2,850 | 7,133 | 40.00% | 23.50% | |
| | B1 | | 1,595 | 815 | 2,410 | 33.80% | 7.90% | |
| | B2 | | 2,693 | 986 | 3,679 | 26.80% | 12.10% | |
| | C1 | | 782 | 189 | 971 | 19.50% | 3.20% | |
| | C2 | | 2,093 | 293 | 2,386 | 12.30% | 7.90% | |
| Dementia (levels) | Independent | 0 | 2,779 | 2,134 | 4,913 | 43.40% | 16.20% | <0.001 |
| | I | | 4,539 | 3,812 | 8,351 | 45.60% | 27.50% | |

| Features | Level | Missing | Stable or Improved (care needs remained the same or decreased) n = 18,659 | Deteriorated (care needs increased) n = 11,728 | Total N = 30,387 | Deterioration Proportion (%) | % in each category | P value |
|---|---|---|---|---|---|---|---|---|
| | IIa | | 1,849 | 1,398 | 3,247 | 43.10% | 10.70% | |
| | IIb | | 3,284 | 2,052 | 5,336 | 38.50% | 17.60% | |
| | IIIa | | 3,331 | 1,550 | 4,881 | 31.80% | 16.10% | |
| | IIIb | | 937 | 354 | 1,291 | 27.40% | 4.20% | |
| | IV | | 1,680 | 406 | 2,086 | 19.50% | 6.90% | |
| | M | | 260 | 22 | 282 | 7.80% | 0.90% | |
| Short-term memory | Unknown | 0 | 5 | 4 | 9 | 44.40% | 0.00% | <0.001 |
| | Capable | | 6,792 | 5,119 | 11,911 | 43.00% | 39.20% | |
| | Problematic | | 11,862 | 6,605 | 18,467 | 35.80% | 60.80% | |
| Cognitive ability | Unknown | 0 | 6 | 5 | 11 | 45.50% | 0.00% | <0.001 |
| | Independent | | 6,819 | 5,389 | 12,208 | 44.10% | 40.20% | |
| | Somewhat challenging | | 4,798 | 3,364 | 8,162 | 41.20% | 26.90% | |
| | Needs assistance | | 4,117 | 2,129 | 6,246 | 34.10% | 20.60% | |
| | Cannot be judged | | 2,919 | 841 | 3,760 | 22.40% | 12.40% | |
| Communication ability | Unknown | 0 | 5 | 8 | 13 | 61.50% | 0.00% | <0.001 |
| | Capable | | 8,460 | 6,689 | 15,149 | 44.20% | 49.90% | |
| | Somewhat challenging | | 4,715 | 3,004 | 7,719 | 38.90% | 25.40% | |
| | Limited to specific requests | | 3,727 | 1,645 | 5,372 | 30.60% | 17.70% | |

| Features | Level | Missing | Stable or Improved (care needs remained the same or decreased) n = 18,659 | Deteriorated (care needs increased) n = 11,728 | Total N = 30,387 | Deterioration Proportion (%) | % in each category | P value |
|---|---|---|---|---|---|---|---|---|
| | Incapable | | 1,752 | 382 | 2,134 | 17.90% | 7.00% | |
| Eating | Unknown | 0 | 33 | 7 | 40 | 17.50% | 0.10% | <0.001 |
| | Independent | | 16,267 | 11,336 | 27,603 | 41.10% | 90.80% | |
| | Fully dependent | | 2,359 | 385 | 2,744 | 14.00% | 9.00% | |
| BPSD | Unknown | 0 | 3,551 | 2,141 | 5,692 | 37.60% | 18.70% | <0.001 |
| | Present | | 4,063 | 2,067 | 6,130 | 33.70% | 20.20% | |
| | Absent | | 11,045 | 7,520 | 18,565 | 40.50% | 61.10% | |
| BPSD | Others | 0 | 392 | 223 | 615 | 36.30% | 2.00% | <0.001 |
| | Filthy behaviors | | 99 | 31 | 130 | 23.80% | 0.40% | |
| | Refusal to care | | 375 | 140 | 515 | 27.20% | 1.70% | |
| | Delusion | | 980 | 573 | 1,553 | 36.90% | 5.10% | |
| | Visual/auditory hallucination | | 15,563 | 10,185 | 25,748 | 39.60% | 84.70% | |
| | Wandering | | 169 | 88 | 257 | 34.20% | 0.80% | |
| | Sexual disinhibition | | 1 | 5 | 6 | 83.30% | 0.00% | |
| | Day–night reversal | | 526 | 185 | 711 | 26.00% | 2.30% | |
| | Assaulting | | 30 | 11 | 41 | 26.80% | 0.10% | |
| | Violent language | | 378 | 149 | 527 | 28.30% | 1.70% | |
| | Mismanagement of fire | | 125 | 130 | 255 | 51.00% | 0.80% | |

| Features | Level | Missing | Stable or Improved (care needs remained the same or decreased) n = 18,659 | Deteriorated (care needs increased) n = 11,728 | Total N = 30,387 | Deterioration Proportion (%) | % in each category | P value |
|---|---|---|---|---|---|---|---|---|
| | Pica behaviors | | 21 | 8 | 29 | 27.60% | 0.10% | |
| Paralysis in limbs | Others | 0 | 275 | 260 | 535 | 48.60% | 1.80% | <0.001 |
| | Right_upper_limb | | 1,282 | 838 | 2,120 | 39.50% | 7.00% | |
| | Right_lower_limb | | 349 | 321 | 670 | 47.90% | 2.20% | |
| | Left_upper_limb | | 3,431 | 1,423 | 4,854 | 29.30% | 16.00% | |
| | Left_lower_limb | | 8,533 | 5,360 | 13,893 | 38.60% | 45.70% | |
| | No_paralysis | | 4,789 | 3,526 | 8,315 | 42.40% | 27.40% | |
| Joint movement without restriction | Others | 0 | 1,507 | 1,109 | 2,616 | 42.40% | 8.60% | <0.001 |
| | Hips | | 1,442 | 787 | 2,229 | 35.30% | 7.30% | |
| | Shoulders | | 3,912 | 1941 | 5,853 | 33.20% | 19.30% | |
| | Knees | | 3,655 | 2,629 | 6,284 | 41.80% | 20.70% | |
| | No restrictions | | 8,143 | 5,262 | 13,405 | 39.30% | 44.10% | |
| Medical treatment in the last 2 weeks_Intravenous therapy | Skin disease treatment | 0 | 341 | 73 | 414 | 17.60% | 1.40% | <0.001 |
| | Catheter | | 371 | 83 | 454 | 18.30% | 1.50% | |
| | Stoma treatment | | 115 | 23 | 138 | 16.70% | 0.50% | |
| | Monitoring | | 36 | 18 | 54 | 33.30% | 0.20% | |
| | Respirator | | 8 | 3 | 11 | 27.30% | 0.00% | |

| Features | Level | Missing | Stable or Improved (care needs remained the same or decreased) n = 18,659 | Deteriorated (care needs increased) n = 11,728 | Total N = 30,387 | Deterioration Proportion (%) | % in each category | P value |
|---|---|---|---|---|---|---|---|---|
| | Central venous nutrition | | 87 | 4 | 91 | 4.40% | 0.30% | |
| | Tracheostomy | | 26 | | 26 | 0.00% | 0.10% | |
| | Infusion management | | 16,559 | 11,229 | 27,788 | 40.40% | 91.40% | |
| | Care for pain | | 81 | 52 | 133 | 39.10% | 0.40% | |
| | Tube feeding | | 484 | 59 | 543 | 10.90% | 1.80% | |
| | Dialysis | | 180 | 72 | 252 | 28.60% | 0.80% | |
| | Oxygen treatment | | 371 | 112 | 483 | 23.20% | 1.60% | |

Table B.3. Accuracy and Model Fitness Results of Care Needs Level Predictive Model in Three Years.

| Model | Classes | Accuracy (Mean ± SD), % |
|---|---|---|
| GBDT | 4 | 74.64 ± 0.04 |
| GBDT | 9 | 53.87 ± 0.55 |
| Logistic Regression | 9 | 41.52 ± 7.91 |

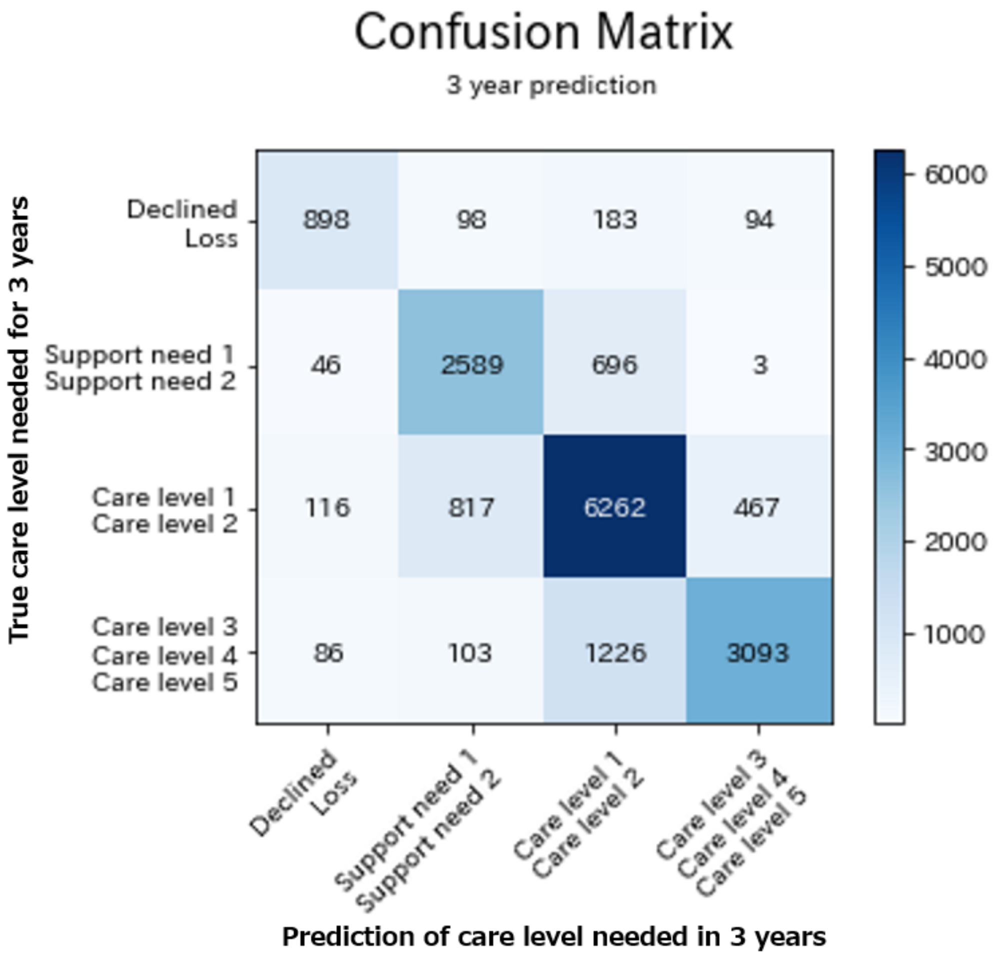

Fig. B.1. Confusion Matrix of the Four-Category Predictive Model.

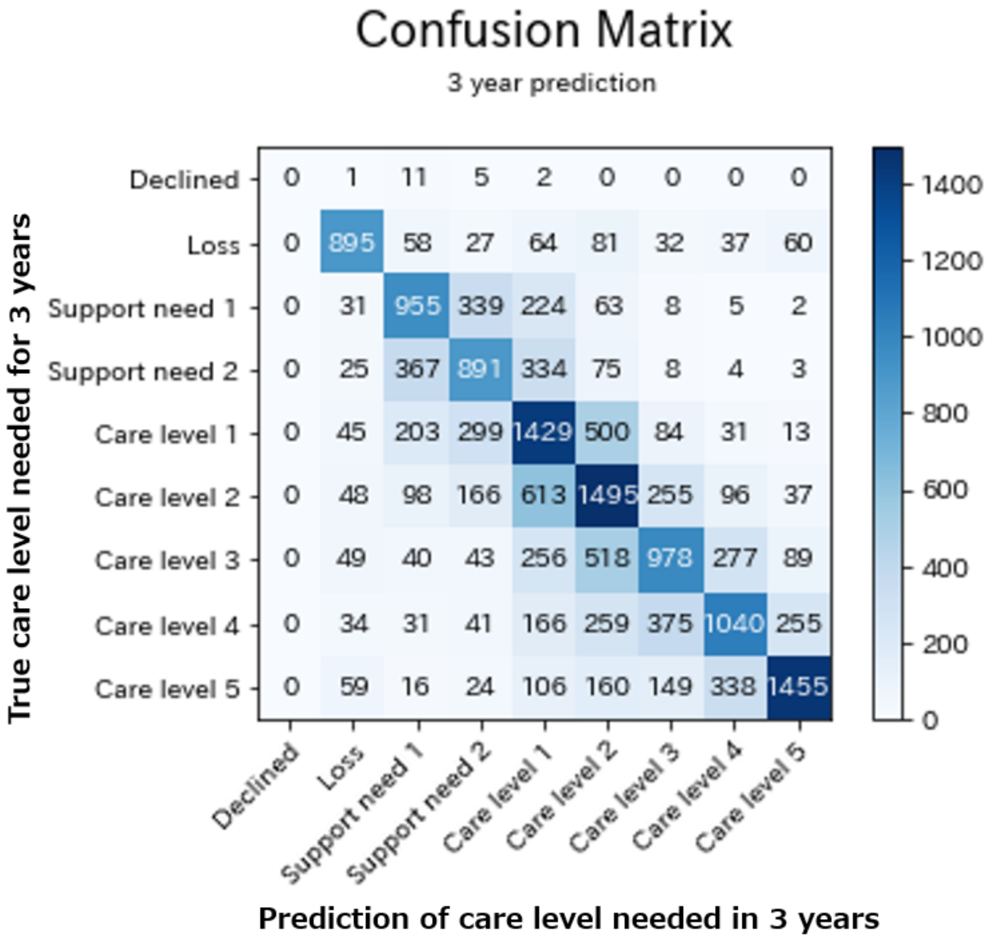

Fig. B.2. Confusion Matrix of the Nine-Category Predictive Model.

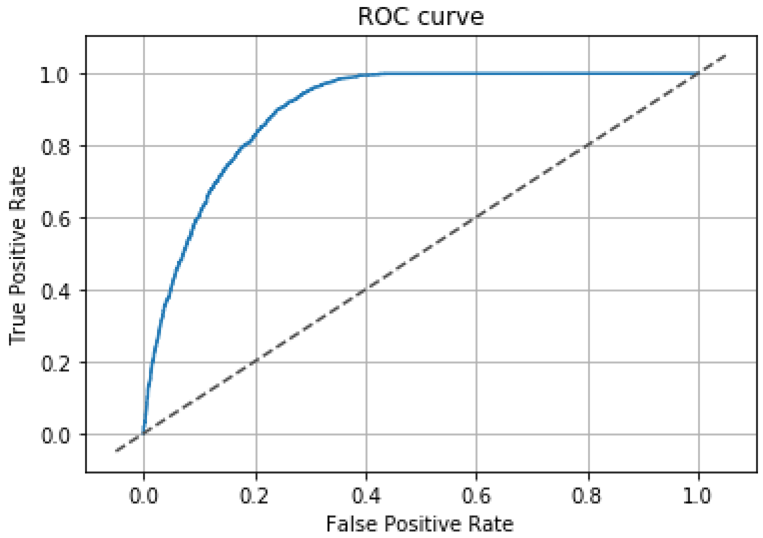

Fig. B.3. ROC Curve of the Binary Gradient Boosting Decision Tree Model.

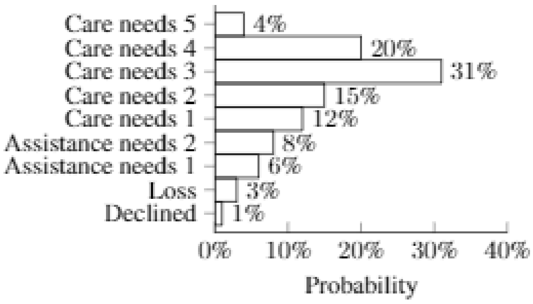


Fig. B.4. Probability distribution of an insureds' predictions.
