## Appendix C Evaluated methods for "Development and validation of gradient boosting decision tree models for predicting care needs using a long-term care database in Japan"

### Evaluated methods

Accuracies, precisions, recalls, F1 scores, area under the receiver-operator curve (AUCs)

The accuracy is the percentage of true positives and true negatives in the total data. Precision refers to the percentage of true positives over true positives and false negatives. The recall refers to the percentage of true positives over true positives and false negatives.

Accuracy = $\frac{TP+TN}{TP+TN+FP+FN}$

Precision = $\frac{TP}{TP+FP}$

Recall = $\frac{TP}{TP+FN}$

F1 score = $2 * \frac{Precision*Recall}{Precision+Recall}$

5-fold cross-validation

To decrease bias in the results, 5-fold cross-validation was conducted. In k-fold cross-validation, the dataset was split into k subsets. The k-1 subsets are used to train a model, and the remaining subset is used for testing the model. This process is repeated k times, using different subset for testing.

Shapley Additive exPlanations (SHAP)

Applying the Shapley values in game theory, SHAP enables each feature's marginal contribution to a prediction to be additively calculated, as illustrate below. If we only had three features, "age," "gender," and "body weight," the SHAP value of age is calculated by aggregating the marginal contributions of elements in coalitions that feature "age," as follows:

$$\begin{aligned} \text{SHAP}_{\text{AGE}}(x_0) = {} & w_1 * \text{MarginalContribution}_{\text{age, \{Age\}}}(x_0) + \\ & w_2 * \text{MarginalContribution}_{\text{age, \{Age, Gender\}}}(x_0) + \\ & w_3 * \text{MarginalContribution}_{\text{age, \{Age, BodyWeight\}}}(x_0) + \\ & w_4 * \text{MarginalContribution}_{\text{age, \{Age, Gender, BodyWeight\}}}(x_0), \end{aligned}$$

where $w_1 + w_2 + w_3 + w_4 = 1$.

GBDT

A single decision tree within the GBDT is a greedy algorithm, and only one feature is selected as a leaf at each iteration. So long as the tree is not designed to hold all features as a leaf, the other multicollinear feature(s) should not be selected.

Several important terminologies related to GBDT should be reviewed. The *nodes* of a tree represent features (attributes), whereas *branches* connect nodes and represent decisions (rules) and *leaves* represent outcomes. *Splits* denote the process of building a tree, whereby the nodes are split based on features and rules and branch out to leaves. *Information gain* is the measure of how meaningful each feature is before and after the node is split—i.e., how much uncertainty is reduced by choosing the feature as a note to split. Finally, *ensemble learning* combines several predictive models (trees) to make a final prediction.

The learning process starts from randomly generating trees, also known as weak learners, and growing the trees by using different features and conditions to make a binary split. The attributes with the highest information gain are chosen until all nodes are split[1]. A single decision tree is appropriate for non-linear dependencies; however, one with a large dataset is usually prone to over-fit[2], which generally results on poorly generalized predictions. GBDT overcomes this problem by using an ensemble learning technique called gradient boosting. A gradient boosting algorithm optimizes the loss function for each tree such that each tree generation pays attention to misclassified trees and their losses from the previous iterations and tries to minimize the loss[3].


1. Sharma H, Kumar S. A survey on decision tree algorithms of classification in data mining. Int J Sci Res. 2016;5(4):2094-2097. https://www.researchgate.net/profile/Sunil_Kumar603/publication/324941161_A_Survey_on_Decision_Tree_Algorithms_of_Classification_in_Data_Mining/links/5aebdfe6a6fdcc8508b6e8bb/A-Survey-on-Decision-Tree-Algorithms-of-Classification-in-Data-Mining.pdf. Accessed September 26, 2019.
2. Horning N. Introduction to decision trees and random forests. whrc.org. http://whrc.org/wp-content/uploads/2016/02/Horning_DecisionTrees_RandomForest.pdf. Accessed September 26, 2019.
3. Schapire RE. The Boosting Approach to Machine Learning: An Overview. In Denison DD, Hansen MH, Holmes CC, Mallick B, Yu B, eds. Nonlinear Estimation and Classification. New York, NY: Springer; 2003:149-171. doi:10.1007/978-0-387-21579-2_9.
